# Description, Development and Application of the Integrated Transport and Health Impact Modelling Tool for Global Cities (ITHIM-Global)

**DOI:** 10.1101/2024.12.11.24318676

**Authors:** Haneen Khreis, Ali Abbas, Daniel Gil-Sanchez, Lambed Tatah, Anna Schroeder, Rahul Goel, Christian Brand, Rob Johnson, James Woodcock

**Affiliations:** MRC Epidemiology Unit, University of Cambridge; Texas A&M Transportation Institute, Texas A&M University System; Transport and Digital Development, World Bank Group, Washington DC; Transportation Research and Injury Prevention Centre, Indian Institute of Technology Delhi, India; UK Energy Research Centre, University of Oxford; Transport Studies Unit, School of Geography and the Environment, University of Oxford; School of Public Health, Faculty of Medicine, Imperial College London

**Keywords:** Model development, Health Impact Assessment Tool, Air Pollution, CO_2_ Emissions, Traffic Incident, Physical Activity

## Abstract

The Integrated Transport and Health Impact Modelling Tool for Global Cities (ITHIM-Global) was developed as an open-source tool to assess the impacts of transport mode shifts on public health and the environment in low- and middle-income countries (LMICs) where the need for such assessment is urgent and rising. The model simulates the impacts on all-cause and cause-specific mortality and years of life lost outcomes through a multi-pathway framework, specifically including physical activity, air pollution exposure, and road traffic fatalities. In addition, the model estimates changes in carbon dioxide emissions resulting from these mode shifts. ITHIM-Global employs a quasi-microsimulation approach that enables individualized exposure estimates by age, sex, and activity levels and utilises up-to-date exposure and dose-response functions that account for the non-linear relationship between physical activity levels and health. To demonstrate its functionality, document its strengths and weaknesses, and aid users, the model was applied to Bogota, Colombia, using three hypothetical scenarios, each shifting 5% of trips to one of bus, car, or cycling. Results indicate that increasing public or active transport modes enhances public health, primarily driven by gains in physical activity and to a lesser extent by reductions in traffic-related emissions. Conversely, a shift to car usage worsens health outcomes due to decreased physical activity and increased pollution exposure. ITHIM-Global serves as a flexible, detailed Health Impact Assessment (HIA) tool adaptable to urban areas across LMICs and this article delves into underlying assumptions and their influence to advance generalisable lessons on risk assessment and model development. The modular structure of ITHIM-Global, its covering of individual and population-level analyses, and extensive documentation provide an accessible, evidence-based approach for city planners and practitioners to optimize transport policies that support public health and mitigate environmental harm.

**Graphical Abstract:** 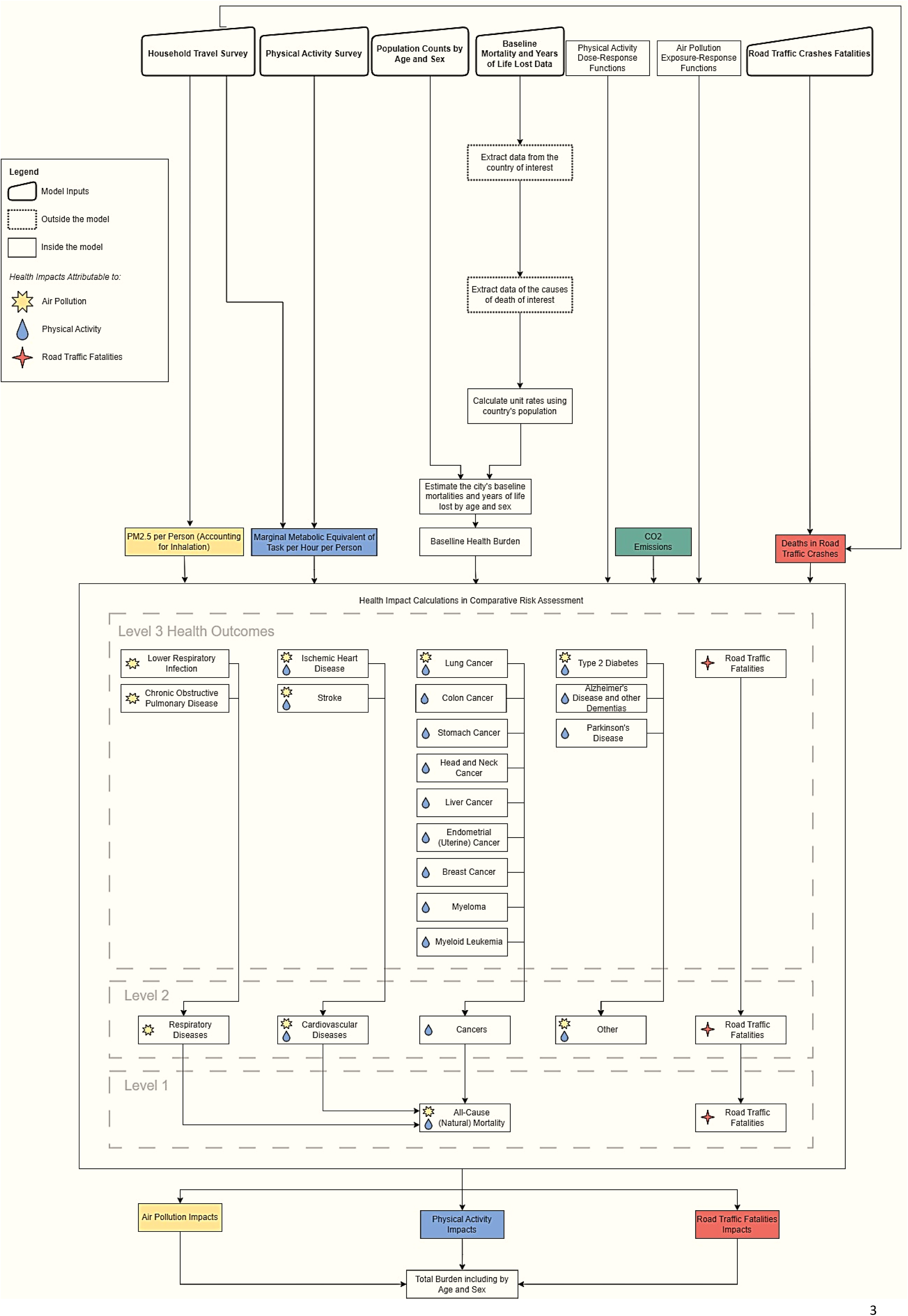

**Highlights:**

- We describe the Integrated Transport and Health Impact Modelling Tool for Global cities (ITHIM- Global), designed for low- and middle-income country (LMIC) cities.
- We apply ITHIM-Global to Bogotá, simulating impacts from three mode shift scenarios with a 5- percentage point increase in car, cycling, and bus shares.
- We simulate the impacts on physical activity, air pollution, road traffic fatalities, and attributable mortalities and years of life lost.
- We describe in detail the model’s elements to aid future uptake, development, and extension to other cities. We discuss strengths, weaknesses, and underlying assumptions to advance generalisable lessons on risk assessment and model development.

## 1. INTRODUCTION

The ways in which people travel around cities impact health through many interacting pathways (Glazener et al., 2021). While detrimental exposures and activities are often concentrated in cities, cities also exhibit adaptability and action agility, presenting opportunities for targeted transport policies (Khreis et al., 2023). By 2050, approximately two-thirds of the global population, close to 7 billion individuals, will live in cities and urban areas (Ritchie & Roser, 2018). Most of this growth is expected in Africa, including the Democratic Republic of Congo, Egypt, Ethiopia, Nigeria, and the United Republic of Tanzania (United Nations Department of Economic and Social Affairs, 2022).

Quantifying the environmental and health impacts of alternative transport policies or scenarios can provide decision-makers and other stakeholders with the evidence needed to consider, champion, and justify action. There is a strong epidemiological evidence-base associating transport-related air pollution and physical inactivity with increased mortality and morbidity (Health Effects Institute, 2022; Garcia et al., 2023). This evidence-base lays the foundation for quantitative Health Impact Assessments (HIA); which can numerically estimate the health impacts of alternative policies or scenarios from those pathways.

Most HIA models remain overly simplistic, often using counterfactual exposures rather than simulating the cascade of changes originating from alterations in transport activities (the source), exposures, and, ultimately, health impacts. Specifically, studies often assume a reduction of air pollution to an “ideal” or recommended level, such as World Health Organization (WHO) guidelines. Such studies frequently fail to elucidate the mechanisms through which such reductions could be achieved or how alterations in transport practices aimed at pollution reduction can give rise to interactions across other pathways. For example, a shift from private vehicles to active transport modes (e.g., cycling/walking) might result in increased pollution inhalation and elevated road injury or fatality risks for active travellers. Simultaneously, this shift enhances physical activity levels among active travellers and potentially decreases ambient air pollution for other commuters and residents.

Quantitative HIAs have generally been applied in the Global North, with a paucity of studies in low- and middle-income countries (LMIC) where the need is most critical (Thondoo et al., 2022). The effective replication, application, expansion, and development of HIAs to LMICs face impediments stemming from the scarcity of data and the absence of open-access, well-documented modelling pipelines configured to data available in those countries. This challenge is compounded by the absence of clear instructions for models’ utilization and transparent documentation of models’ assumptions.

Here, we describe a HIA model, the Integrated Transport and Health Impact Modelling Tool (ITHIM), along with its most recent configurations to cater to applications in cities of LMICs denoted as ITHIM- Global. A recent review identified 87 HIAs of transport since 2015 (Mizdrak et al., 2023). The most used model was ITHIM, followed by the WHO’s Health Economic Assessment of Transport (HEAT) tool for walking and cycling (Kahlmeier et al., 2017).

This article provides a comprehensive view of ITHIM-Global, explaining its intended purpose and capabilities, its constituent elements and modules, requisite input data and their format, and modifications to inputs during output generation, as well as underlying computations, estimations, and assumptions. Currently, ITHIM-Global models health impacts through three pathways from transport to health: physical activity, air pollution, and road traffic fatalities, with each pathway’s assessment termed as a “module” in this article. Additionally, the model assesses transport-related CO_2_ emissions resulting from modified passenger travel patterns.

To enhance clarity, the model’s components are systematically illustrated through an exemplary case study in Bogotá where we simulate an archetypal mode shift scenario with a 5% increase in mode shares for three target modes:

1. Buses,
2. Cars (private vehicles),
3. Cycling

ITHIM in its different forms has been previously described in various papers. Of most relevance here is the description and application of the model in its original Excel spreadsheet format in England and Wales (Woodcock et al., 2013), in Analytica in São Paulo (de Sá et al., 2017), in R, independently developed in the US, and applied in California (Maizlish, 2019), and most recently, in R, applied in Accra, Ghana (Garcia et al., 2021). These papers represent developments and modifications for specific use cases. The implementation presented here builds upon these and subsequent methods development described in this article. This article represents the first account of the ITHIM-Global model and its various elements to facilitate its future uptake, further developments, and extensions for LMICs.

## 2. MODEL DESCRIPTION

### 2.1. Model Overview and Elements

ITHIM-Global is implemented in the R language and accessible on GitHub: https://github.com/ITHIM/ithim-r. It is designed to evaluate, at the city level, the health impacts resulting from changes in passenger (not freight) travel, considering physical activity, air pollution, and road traffic crashes. The preprocessing stage establishes a baseline population derived from individual-level travel surveys and other relevant data. Subsequently, the model utilises this population to simulate alterations in passenger travel patterns by modifying the mode share of trips, estimating consequential changes in individual-level physical activity, air pollution exposure, fatalities from road traffic crashes, and attributable health impacts. Each step within the model entails definitions, inputs, and outputs, described here, and coded in a modifiable manner. The model employs a comparative risk assessment approach (Ezzati, 2008), with a quasi-microsimulation exposure assessment which uses the created baseline population to assign individual exposures (Scarborough et al., 2014). The health impacts from a given scenario are determined by estimating mortalities and years of life lost (YLL) attributable to each pathway separately, alongside an assessment of the combined multi-pathway impact. Additionally, the model estimates CO_2_ emission changes.

There are two operational modes for the model: the first assumes fixed input parameters, while the second involves a Monte-Carlo analysis creating distributions for input parameters and running the model multiple times, each utilizing different samples from these distributions. This article describes the fixed modality, with a forthcoming article to detail the second modality. Figure 1 shows the model’s elements and how they interact. The three modules, air pollution, physical activity, and road traffic fatalities, are denoted in different colours. Some elements are shared across the modules.

**Figure 1.**
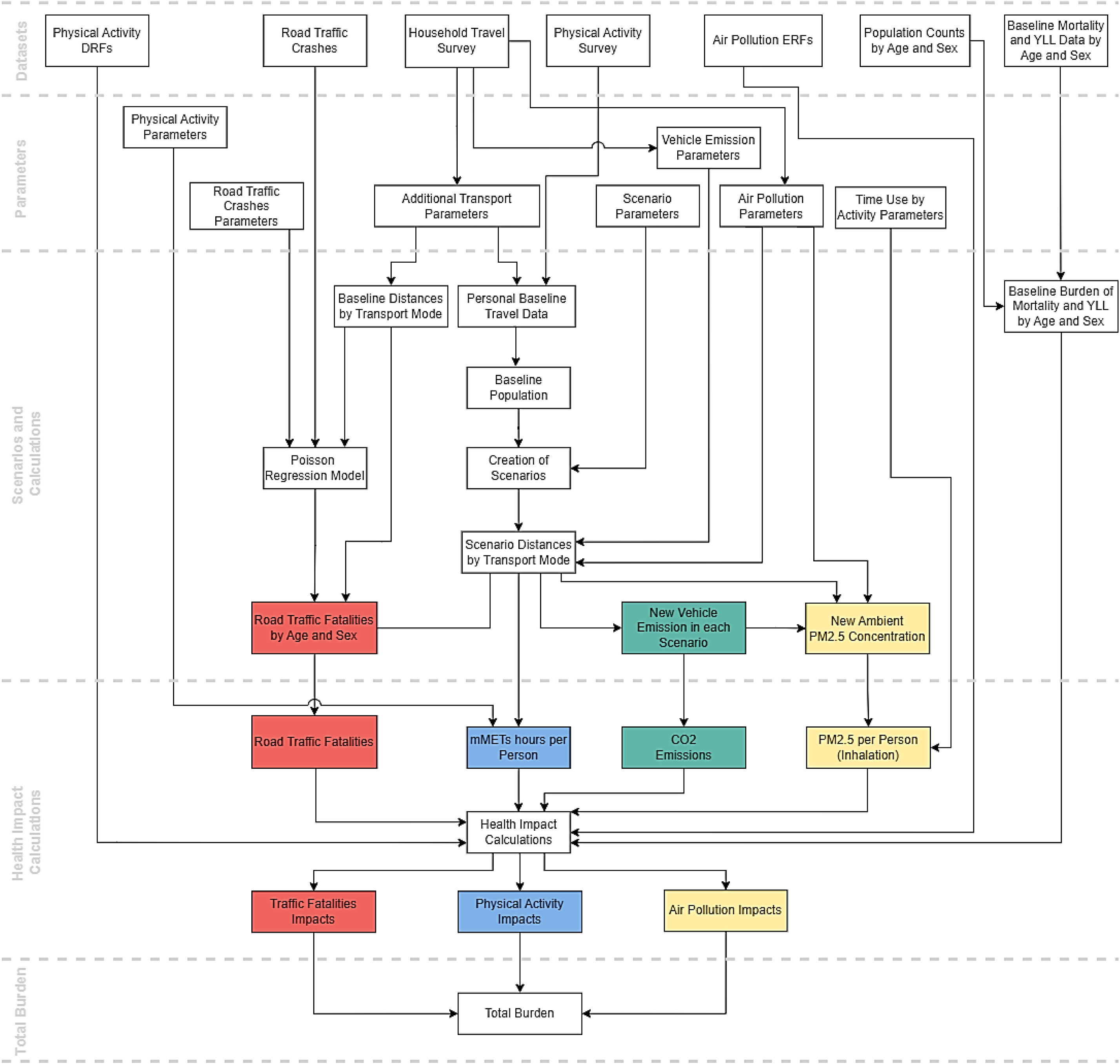
Simplified ITHIM-Global Workflow Denoting the Underlying Elements of the Three Modules: Road Traffic Fatalities (Red), Physical Activity (Blue), and Air Pollution (Yellow)

### 2.2. Developments over Previous Versions

ITHIM-Global improves upon previous implementations by (1) using a quasi-microsimulation approach to represent travel patterns, physical activity and air pollution exposures at an individual level; (2) updating and extending exposure-response functions (ERFs) for air pollution for all-cause and cause-specific mortalities; (3) more robustly estimating exposures during travel and non-travel activities accounting for sex- and age-specific inhalation rates; (4) updating and extending dose-response functions (DRFs) for physical activity using harmonised methods across more endpoints and including all-cause and cause-specific mortalities; (5) changes to the statistical model estimating traffic fatalities and (6) the documentation and packaging of the model and code in open-access resources.

The next sections take the reader through the overall model. We describe the required input data, their format and transformations, and the underlying computations, estimations, and assumptions of the three modules. We provide information on running the model and producing outputs, including the interpretation and verification of intermediate and final outputs, using data from Bogotá as an example.

## 3. MODEL BREAKDOWN

### 3.1. Baseline Travel Data

Travel data are central to ITHIM-Global and are featured at several stages in the modelling process including the creation of the baseline population and counterfactual scenarios. The baseline travel data comprise information on individuals and their trip characteristics and travel distances. The primary data source is city-specific household travel surveys, complemented by estimates from reports that provide additional transport parameters.

#### 3.1.1. Household Travel Survey Data, Pre-Processing, and Harmonization

Household travel surveys provide most of the baseline travel data and serve as the basis for creating cities’ baseline populations. Thus, the selected travel survey should provide information on each participant’s age, sex, and trips.

Trip information includes the number of trips and stages, purpose of travel, modes, and travel durations. Surveys typically describe trips at the main mode level or sometimes provide separate records on the stages that make a trip. The main mode is typically defined using the longest duration or, ideally when available distance. A stage is completed within a trip when an individual transfers from one mode to another. For example, a bus trip typically includes three stages: walking to and from the bus stop and the stage travelled on the bus. Sometimes, only limited information is provided about stages, such as stage mode, with no information on distances or durations. The ideal survey contains separate files on trip and stage characteristics. Many cities, including in LMICs, have travel surveys. For Bogotá, we accessed the 2019 survey from the <u>Integrated Information System on Regional Urban Mobility</u>.

Typically, substantial pre-processing and harmonization are required to transform raw survey data into a format suitable for ITHIM-Global.

Travel survey data are often provided in three files: one containing information about the households, one about people living in households, and another for trips they reported during the survey. A fourth dataset would contain information about trip stages. There typically is a survey report describing the survey and geographical coverage. Travel surveys may just cover a single city or include neighbouring areas. The survey’s geographical boundaries must be established because travel data must spatially match other datasets used in ITHIM-Global, e.g., population, crashes and vehicle emissions. If the survey is not confined to city boundaries, a data subset should be taken to keep only the participants within the city of interest. Assessment of data quality, processing, and harmonisation can then proceed as follows.

We first replicate the original summary statistics in the report to ensure our understanding of trips, stages, modes, and trip distances/duration align with the survey report. For example, some surveys report all trip stages as independent trips, not segments within a trip, or include waiting time at stations as part of trip durations. Pre-processing then combines variables from different datasets into a single dataset with trip and participant information. Our unit of analysis is either stages or trips. Data harmonisation extracts stage information from trip-level data and harmonises transport modes and duration definitions (e.g., excluding waiting time for bus). If detailed stage-level data are unavailable or only partially available, more stage information may be derived from trip data, but with caveats. Often, only stage mode is available, not stage duration. In that case, though not recommended, trips may be used as the smallest unit of analysis with the limitation of losing stage-level information. However, heuristics can be used to derive missing stage durations. In ITHIM-Global, the user has the option to set the “`add_walk_to_pt_trips`” input parameter to “`true`” and assign a walking stage mode and duration to all bus trips that do not already have a walking stage. The duration of walking to/from the bus stops may be taken from surveys of similar settings (Goel et al., 2022). So, walking stages are automatically added to those bus trips without walking stages during an ITHIM-Global run if the trip duration is long enough (see Section “3.1.2. Additional Required Transport Input Parameters”). Data on walking to/from the bus are particularly important as they directly impact pathways, e.g., the amount of physical activity from walking and exposure by mode. In Bogotá, the travel survey had detailed stage-level data. Surveys often use local names for transport modes that do not match the ones used in the model. Names can be mapped based on pre-established categories in Table S1.

The processed data are then quality assessed by comparing the following estimates against estimates from similar cities to check their plausibility: number of trips per capita, total travel duration per capita, and mode shares. The quality of travel surveys is affected by many factors, such as response and recall rates, proxy responses, soft refusal, representativeness of the population, duration limitations e.g. covering unrepresentative one-day travel behaviour, lack of spatial coverage/missing geographies, the absence of some modes which are a minority and/or which are sometimes categorised as “other”, especially motorcycle or cycling, and the absence of occupation-related travel. Some of these limitations are not dealt with in our model. Investigations of poor data quality should consider these factors and where applicable use methods to adjust survey data such as imputing motorcycle or cycling trips or subtracting waiting time from trip durations where durations are too long and judged to include time waiting for the bus. We assessed and judged that the Bogotá survey was of sufficient quality. The exact variables required for ITHIM-Global from the survey are outlined in Table S2 of the Supplementary Material. Pre-processing and harmonisation steps are depicted in Figure 2.

**Figure 2.**
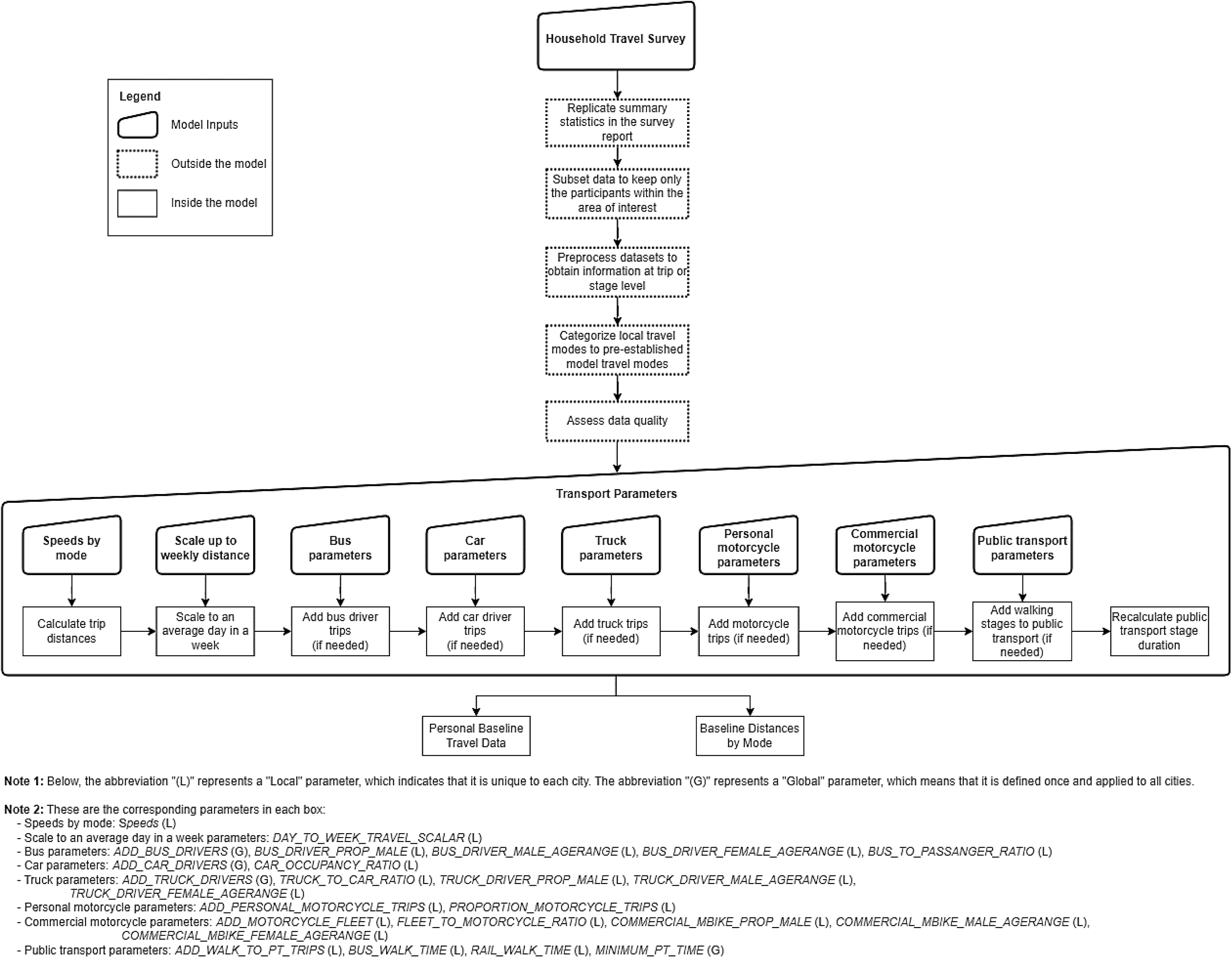
Flowchart Outlining Pre-Processing and Harmonisation of Household Travel Surveys

#### 3.1.2. Additional Required Transport Input Parameters

Travel surveys provide information about person travel behaviour but less on corresponding vehicles, such as speed and occupancy, and exclude occupation-related travellers (couriers, taxi drivers, lorry drivers). These data are required to complete information on person and vehicle travel, and different imputation methods can be considered. The additional transport-related parameters need to be defined locally for each city and are illustrated in Table S3 for Bogotá, while global parameters, which apply to all cities, are in Table S4. Local parameters include (1) mode speed, used to calculate trip distance; (2) multiple parameters to define walking to the bus; (3) personal motorcycle trips where not recorded in the survey but are expected; (4) commercial motorcycle trips; (5) truck trips; (6) bus and car occupancy ratios to estimate vehicle distance travelled and (7) a temporal scaling parameter. The source or derivation of, and underlying assumptions for, each variable in Bogotá are presented in Section S3 in the Supplementary Material.

### 3.2. Creating the Baseline Population

We refer to the population from the travel survey, matched to a physical activity (PA) survey, as the baseline population. The baseline population provides the individuals who are assigned travel characteristics and contains demographics and non-travel PA data. In this baseline population, each individual represents other individuals in the study area based on variables used in the selection of survey participants, such as location, car ownership, and socioeconomic status.

To create the baseline population, we use individuals’ identifiers, age, and sex, from the household travel survey prepared in the previous section, and non-travel PA derived in a later section. We assume that the survey’s respondents are representative of the city’s demographics. Users should compare travel survey demographics with those from the most recent census or other official datasets. The travel survey for Bogotá included 52,174 participants, ranging in age from 0 to 110, with a mean age of 37.2 years; 53% of the participants were women. According to the 2019 Bogotá census, the city had a population of 7.6 million, with 52% being women and included ages ranging from 0 to 100 years. Table S5 shows a relatively similar 5-year age band population distribution when comparing the 2019 census with the travel survey. Table S6 demonstrates the variables of the baseline population.

If the travel survey is unrepresentative, users can utilise participant weights, which are often included with the travel survey and show how many people in the general population of the studied area are represented by an individual respondent. Expanding the survey by weight means creating new sets of participant identifiers which take on the unweighted participant’s demographics. The weighted population can then be used to build the baseline population. Census or other data can be used to create weights if survey weights are not available.

Currently, ITHIM-Global focuses on populations aged 15-69 years in cities of the global south, where the population age structure skews younger. For example, the median age in 2021 in Colombia was 30.8, compared to 37.7 and 39.6 in the US and UK, respectively (Ritchie & Roser, 2019). We excluded younger children because they are not well represented in many travel surveys and because our mortality-based model is not well suited for capturing the effects of air pollution or physical activity in children, although they represent a significant proportion of LMIC populations. Using the model for adults older than 69 years needs careful consideration. The method we use to re-allocate trip modes might generate implausibly high levels of walking and cycling in this age group, which can lead to exaggerated PA benefits as there is an exponential relationship between age and mortality.

Individuals’ non-travel PA is required for the PA health impact calculation. We use non-occupational PA. To assign non-occupational PA to individuals, we matched individuals in the travel survey with those in a PA survey (see Section “3.7. Module 2: Physical Activity Pathway”). The matching of individuals can be done in various ways. ITHIM-Global samples once by matching the age group (five-year age band) and sex (male or female) between the two data sources. We ensured that the proportion of individuals without any background physical activity (i.e., marginal MET = zero) is maintained by age-band and sex. The baseline population individuals are assigned different travel characteristics under different scenarios, and the resulting datasets feed into the distance calculations, road traffic fatalities, physical activity, and air pollution modules. We use the baseline population whenever we are interested in person-level calculations such as distance travelled by mode per person, physical activity per person, air pollution exposure per person and injury risk.

Our approach, utilising the travel survey to build the baseline population, sits between a fully macro-level population, which uses summary statistics, such as mean age and sex distribution, and a full-scale spatial micro-simulation of the city’s entire population (Grieves, 2014). We use baseline population individuals to generate what-if scenarios and estimate individual-level exposures as in a micro- simulation, but we ultimately estimate health impacts for aggregated age groups, making the approach quasi-micro-simulation.

### 3.3. Creating Scenarios

Various methods exist for specifying counterfactual travel behaviour scenarios. In ITHIM-Global, we adopt a straightforward approach where the trip is considered constant, and only the mode can undergo changes governed by stochastic rules contingent upon trip distance. This approach is simplistic, particularly in neglecting interactions such as those between mode and destination choice, which can be captured using a full agent-based travel behaviour model. Nevertheless, there is utility in isolating and examining changes from specific elements, such as mode choice and share.

For our application in Bogotá, we define scenarios by increasing the trip share of target modes: cars, buses, and bicycles by five (5) percentage points (PP), drawing from baseline trips. Table 1 shows the mode split in the baseline and the three scenarios. In each case, the mode share is increased by 5 PP. For example, in the car scenario, the baseline mode share of 18.1% is increased to 23.1% (18.1+5). In sensitivity analyses, we increase target modes by 1 PP and 10 PP. We use these scenario definitions to balance comparability and realism. Using the same PP increase for each transport mode increases comparability. However, different trips are changed in different scenarios, e.g., in the car scenario, the trips shifted to the target mode Car will be different than the trips shifted to the target mode Bus in the bus scenario. In all cases, trips are randomly re-assigned to the target scenario mode based on distance rules. In Bogotá, the 5 PP (or 1 or 10 PP) increase in the target mode trip share is split across three pre- defined distance categories in ITHIM-Global (0-2km, 2-6km and >6km) using both the mode shares by distance band (Table S8), and the share of total trips by distance band (Table S9), as explained in full in the Supplementary Material.

**Table 1.** Percentage Mode Split Between The Baseline and Three Created Scenarios.

|  | Scenarios |  |  |  |
| --- | --- | --- | --- | --- |
| % Mode split | Baseline | Car | Cycling | Public Transport via Bus |
| Car | 18.1 | 23.1 | 17.1 | 15.3 |
| Bus | 34.8 | 31.2 | 32.9 | 39.8 |
| Cycle | 5.2 | 4.9 | 10.2 | 4.8 |
| Pedestrian | 31.2 | 30.8 | 29.4 | 30.7 |
| Motorcycle | 5.2 | 4.7 | 4.9 | 4.3 |

### 3.4. Baseline Mortality and YLL Data

ITHIM-Global employs a comparative risk assessment approach to estimate health impacts in the form of mortalities and YLL due to scenario-related changes in PA, air pollution and injury risk amongst the baseline population. This approach requires baseline mortality rates (all-cause and cause-specific), YLL, and city-level population counts.

Baseline unit rates for all-cause and cause-specific mortality and YLL can be estimated using the Global Burden of Disease study (GBD) data published by the Institute for Health Metrics and Evaluation on a yearly basis; 2019 was the latest year at the time of analysis (Global Burden of Disease Collaborative Network, 2020). The GBD offers a freely available and standardised dataset that can be used across many countries. Users may choose to compare results using local versus country-level (GBD) data if local data are available.

Estimates of population sizes, baseline mortality numbers (counts) for “all-cause” mortality, and cause-specific mortalities and YLL can be extracted from the GBD at the country level (subnational level, if available). Mortality counts divided by the country’s (subnational spatial unit’s) population counts give the unit rates for outcomes of interest. There may be differences between city and country-level event rates, and adjustments can be made if suitable data are available. Data are extracted in quinquennial age ranges (most resolved) and separated by sex (females, males).

Within ITHIM-Global, the calculated unit rates are used to estimate the city’s baseline mortalities and YLL by multiplying the unit rates by the city-level population counts extracted from the census in the same 5-year age and sex bands. For Bogotá, we used population projections for 2019, the year of the travel survey and GBD, by sex and 5-year age band. Table S10 delineates the GBD and census data items and their required data format for input into ITHIM-Global.

### 3.1. Mortality and YLL Outcomes

Epidemiological studies relate exposures and mortality outcomes at multiple levels: (1) all-cause and cause-specific, including (2) disease groups (e.g., cardiovascular, respiratory, and cancers), and (3) specific diseases (e.g., type 2 diabetes, stroke, chronic obstructive pulmonary disease) mortality and YLL. To elucidate scenarios’ overall and disease-specific impact, we report mortality and YLL outcomes at these three levels, shown in Table S11 and Figure 3.

**Figure 3.**
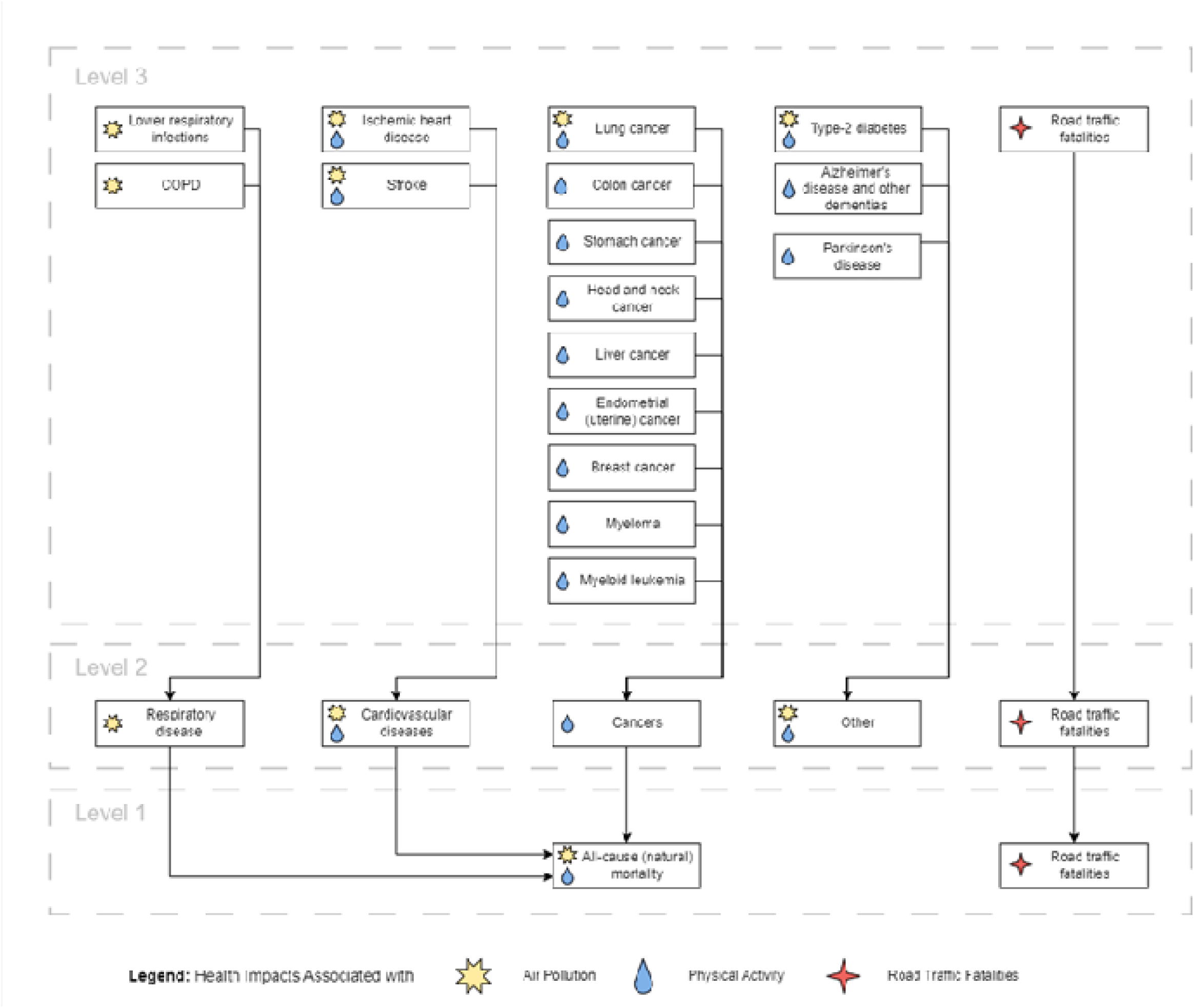
Mortality and YLL Outcomes included in ITHIM-Global and their Levels (1, 2, and 3)

The cause-specific mortalities and YLL we selected were those which have been convincingly associated with physical activity and/or air pollution exposure. The physical activity and air pollution modules include information about the selection of DRFs and ERFs. To estimate the interaction between these pathways, we multiply risk ratios from both pathways and use the resulting risk ratio for a combined interactive impact.

### 3.2. Module 1: Road Traffic Injuries (RTI) Pathway

The RTI module estimates mortality and YLL from fatal road traffic crashes only. Figure 4 illustrates the process of modelling traffic fatalities under created scenarios.

**Figure 4.**
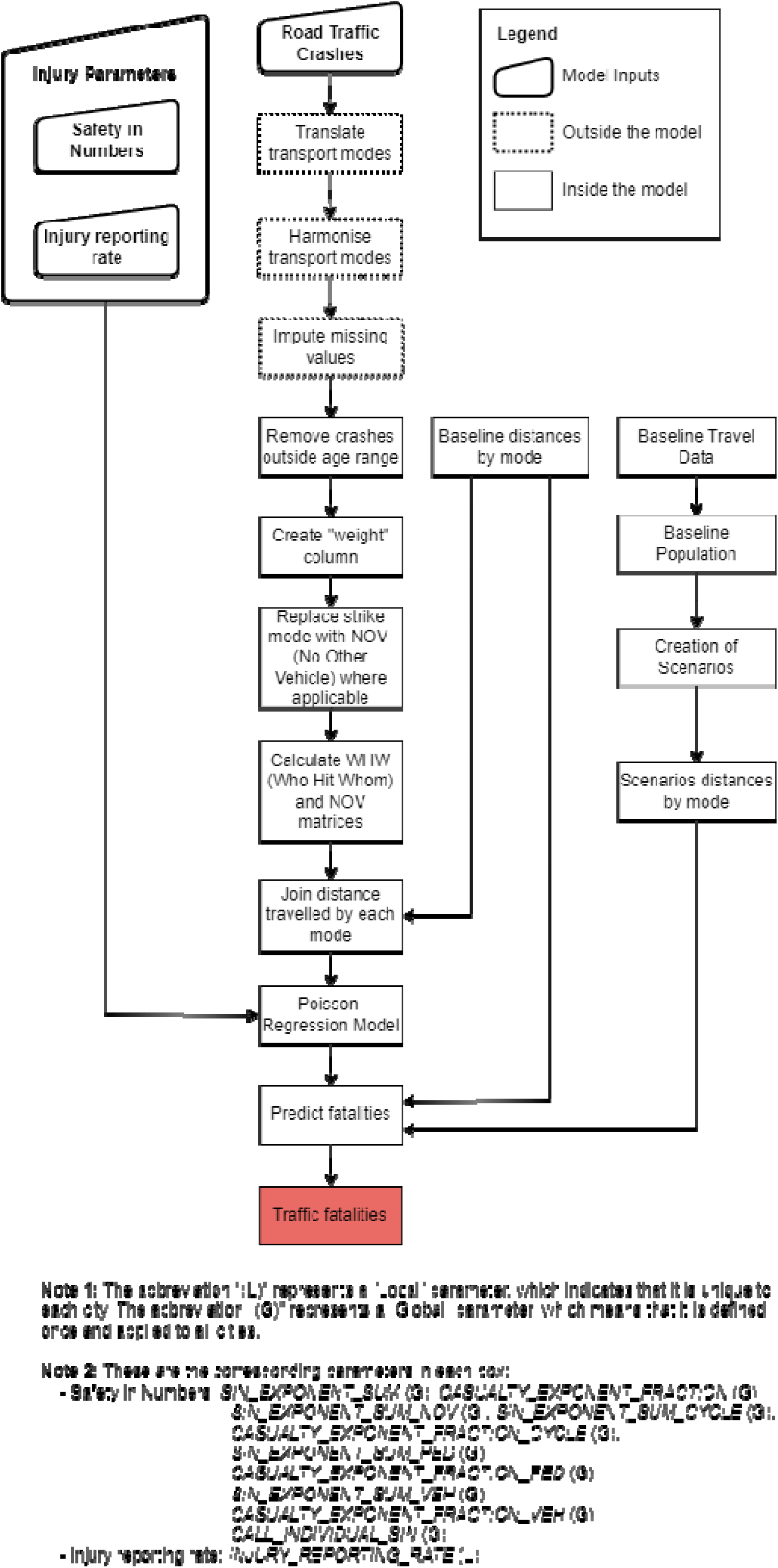
ITHIM-Global Workflow for the RTI Module

The main input for the RTI module is a data frame with rows representing casualties who died in fatal crashes and columns indicating each casualty’s mode of transport (and age and sex if available, as is the case for Bogotá), and the other vehicle(s) involved in the crash (referred to as the strike vehicle). These data come from either police or, less commonly, hospital/mortuary records. A single year’s data may no be representative of longer-term averages, particularly when stratified. Using records from a longer period makes data more stable when stratifying by age, sex, and mode. For Bogotá, we accessed police department data for a five-year period (2015-2019), compiled and shared by the Bogotá Secretary of Mobility in partnership with the World Resources Institute.

The RTI module data frame has one row per fatality. The source data for Bogotá contained information at the crash level. For each fatality in each crash, we added a row with the casualty’s details and assigned the strike vehicle using information from the crash record. When multiple fatalities were recorded for a single crash that involves more than one vehicle, we added casualties to the data frame as if they were from separate crashes. For instance, a crash involving a motorcyclist and a car where both the car occupant and the motorcyclist are killed is equivalent to two separate crashes between a car and motorcyclist where there is one fatality in each crash. The first record has the car occupant as the fatality and the motorcycle as the striking vehicle, and the second record has the motorcyclist as the fatality and the car as the striking vehicle. Where there were more than two parties in a crash record, we selected the largest other vehicle as the strike vehicle. For instance, if a pedestrian was killed in a collision with a car and a truck, we assumed the truck was the strike vehicle. For single-vehicle crashes, we denoted the strike vehicle as “no other vehicle” (NOV), with the exception of pedestrian fatalities. For these pedestrian cases, we used Multivariate Imputation by Chained Equations (MICE) via the mice package in R (Buuren and Groothuis-Oudshoorn, 2011) to impute strike vehicles. We then harmonised the names of transport modes with names in ITHIM-Global (Table S1). Age and sex information was provided for most casualties in the Bogotá’s crash data. Missing data (including age, sex, and mode of transport) were replaced with imputed values again using the mice package. We impute missing values to maximise the number of data points used to fit the crash-fatalities model described later. All these steps happen before the data set is read into ITHIM-Global.

The second input data are the baseline distances by mode, processed in Section 2.1. As discussed, we also added commercial motorcycle trips, truck trips and bus driver trips in the case of Bogotá because inclusion of travel by these vehicles is important to accurately model fatality counts. To find the total distance travelled by all people by bus, we add the bus driver kilometres to the bus passenger kilometres. Note that the car trips found in the travel survey already consist of car drivers and car passengers, so we do not need to add car drivers explicitly. We also multiply the taxi and auto rickshaw passenger distances from the travel surveys by 2 to account for taxi and auto rickshaw drivers. When multiple passengers in taxis or auto rickshaws have been captured in the travel survey, then multiplying the passenger distances by two overestimates the total distance travelled by people (passengers plus driver) in those vehicles. However, both taxis and auto rickshaws spend a significant amount of time driving without any passengers (UITP, 2020), which may counteract this overestimation.

The third set of inputs needed is related to the safety-in-numbers coefficients defined by the user. The safety-in-numbers effect states that the number of crashes tend to grow less than in proportion to traffic volume (Elvik & Goel, 2019). Two sets of safety-in-numbers coefficients are defined for casualty (`casExponent)` and for strike mode distances (`strExponen`t) (Table S12). These coefficients are used in the Poisson regression model to ensure that we include safety–in-numbers effect.

An ITHIM-Global user can either use one coefficient for all casualty modes and one for all strike modes or define separate values for individual modes. The model includes a logical parameter, `call_individual_sin`, which, when set to false, means that one coefficient is used for all casualties and one coefficient for all strike modes. Otherwise, individual casualty and strike mode coefficients are used for vehicles, bicycles, pedestrians, and NOV crashes. For Bogotá, we set `call_individual_sin` to `true`. Instead of inputting these casualty and strike exponents directly, we input the sum of the `casExponent` and `strExponent` as `sin_exponent_sum` and then define a `casualty_exponent_fraction` parameter, which defines the fraction of the `sin_exponent_sum` that is attributed to the `casExponent` parameter. When the model is run for several cities, these exponents apply to all cities to allow comparison across cities. Table S12 documents the safety-in-numbers coefficients for Bogotá, while the `sin_exponent_sum` and `casualty_exponent_fraction` input parameters are given in Table S13.

The final input is an estimate of the fraction of fatalities that we expect are reported at the city level using the `injury_reporting_rate` parameter. We estimated this fraction for Bogotá based on country-level data from WHO (WHO, 2018) that show both recorded and estimated crash fatality numbers for Colombia, giving a national reporting rate of 80% (7,158/8,987). We assume that urban reporting for the capital city is better than rural reporting. Therefore, we halved Colombia’s under- reporting to obtain a value for Bogotá (i.e., we assumed an under-reporting of 10%). As such, we set our `injury_reporting_rate` parameter to 0.9. We recommend that users utilise local data if they have access to city-level under-reporting rates or can estimate these. We recommend estimation and analysis of traffic fatalities burden per 100,000 people, billion km, and 100 million hours, separately by mode at baseline, to investigate data and assumption plausibility.

When the ITHIM-Global model is run, the traffic crash dataset is read in, and crash fatalities outside the age range of 15-69 are removed. If age information is unavailable, we apply the proportion of crash fatalities for the age range of interest from the GBD data to retain crash fatalities for the age group we are modelling. We also assign a ‘weight’ column to denote the number of years for which we have crash data. For Bogotá, this was set to 5 (2015 – 2019).

For crashes where the casualty mode is the same as the strike mode, we set the strike mode to NOV as we assume that risk scales similarly to NOV collisions (see Fatality Prediction Model Details in Section S9.1 of the Supplementary Material). The data are then split into a WHW (who hit whom) part, where both strike and casualty modes are known, and a NOV part, where the strike vehicle is NOV. Both datasets are aggregated by casualty and strike mode (and age and sex where this information exists) to get total fatality counts for each casualty and strike mode (and age and sex category) combination. These fatality counts are then joined with the distance travelled by each strike and casualty mode as calculated during the ITHIM run and by age and sex, if available. Suppose there is age and sex-specific fatality information, but the corresponding mode distances for a specific age and sex category do not exist. In that case, any age and sex information is removed, and fatality counts and distances are aggregated by casualty and strike mode only. These datasets containing total fatality counts and distance information for each casualty and strike mode pair (and age and sex category where applicable) are used to build a fatality prediction model.

Given that traffic fatalities are relatively rare events when considering combinations of modes and age groups and sexes, a Poisson regression model is used. The expected number of crash fatalities for any given combination of casualty mode and strike mode (and age group and sex where available) is estimated using a Poisson regression model fitted to the reported counts and distances travelled (as in, e.g., Miaou, 1994). The specific regression terms are chosen by starting from the form shown in the Supplementary Material, and if this form returns standard errors higher than 10 for any of the coefficients, the model is modified by aggregating age and sex categories.

Using the fitted model, fatality counts for the different scenarios and the baseline scenario are modelled for each casualty, strike mode, age group and sex. If the Poisson model was not set up to take age and sex into account due to missing data, then predictions are based purely on distances travelled by different modes in each age and sex category. Modelling baseline data ensures consistency between baseline and scenario fatality estimates and smoothens out any particularly high or low fatality counts in the raw data, which can be expected for some casualty and strike pair combinations. Table S13 documents the parameters required for the RTI Module.

### 3.3. Module 2: Physical Activity (PA) Pathway

The PA module estimates mortality and YLL from changes in individuals’ PA volumes as their transport-related walking and cycling change across scenarios. To perform estimations, we use (1) module-derived estimates of individuals’ PA volumes in the baseline and counterfactual scenario(s), and (2) DRFs derived from meta-analyses, which associate PA volumes with various health outcomes (Garcia et al., 2023). Elements and parameters of this module, and how they feed into each other are in Figure 5 and Table S14, and described next.

**Figure 5.**
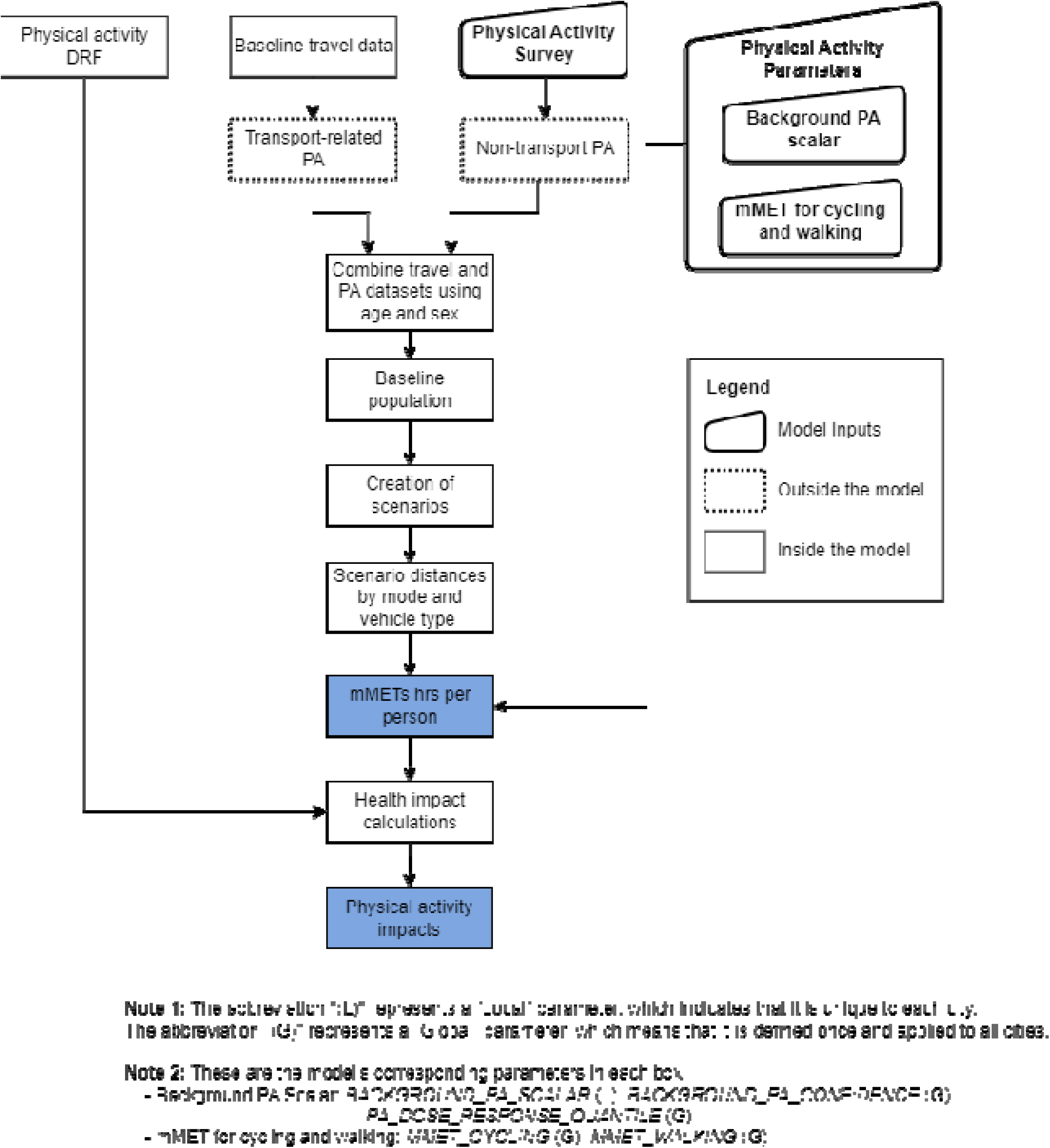
ITHIM-Global Workflow for the Physical Activity Module

PA is often described across four main domains: occupational (as part of paid work), domestic (as part of household chores or domestic tasks), leisure-time (as part of leisure, such as sports and exercise), transport (as part of transport, such as walking and cycling). Sedentary behaviours (sitting or lying down, e.g., when watching television) are physically inactive and detrimental to health, but the harms of sedentary time while travelling are not currently included in the model.

There is good evidence that the relationships between PA and health are non-linear. This implies that relative risk (RR) changes are not linearly proportional to PA changes but depend on background PA (Garcia et al., 2023), with the greatest differences observed at low activity levels. While PA volumes from the four domains are known to affect health and should contribute to background PA, we focused on the health impacts of non-occupational PA domains, which have more robust and consistent supporting epidemiological evidence (i.e., increasing PA increases health benefits). We exclude occupational PA from our background PA because it has a complex and multidimensional relationship with health, with potentially positive or negative health effects depending on the nature, intensity, and duration of the activity, as well as the individual’s overall health status and other lifestyle factors (Clays, 2018). Moreover, occupational PA is often poorly captured in surveys that tend to assign a fixed value to hours worked when the actual intensity of the work will vary over the day and week and within a job type (Pedersen et al., 2016). Therefore, individual PA volumes used in ITHIM-Global comprise transport, leisure and domestic PA. Leisure and domestic PA constitute the non-transport, non-occupational PA, although domestic PA is often not reported as a separate category in PA surveys, so we typically use leisure PA as a proxy for both.

Information on non-transport, non-occupational PA comes from surveys, which typically use the Global Physical Activity Questionnaire (GPAQ), the International Physical Activity Questionnaire (IPAQ), or modified versions of these. We rely on data from questionnaire surveys because they are widely available, as are corresponding RR; they capture data in domains and are easier to integrate with our travel survey compared to the less biased PA measures from objective sources such as accelerometers and wearable fitness trackers. PA surveys are subject to a wide range of biases that limit their validity, including self-reporting, social desirability, recall, question misinterpretation, seasonal variability, and participant fatigue. Future data efforts should consider combining questionnaire and objective data to reduce bias and still capture PA in separate domains. The surveys provide standardised data processing guidelines (GPAQ, 2021; Sjostrom et al. 2005). Typically, the PA volume of each type of activity is obtained by multiplying daily duration, weekly frequency, and PA intensity.

Different PA types, such as walking and cycling, are assigned intensities based on values from the Compendium of Physical Activity (Ainsworth, 2011). When PA is captured in an aggregate manner, i.e., moderate, or vigorous intensities without activity specification, as in GPAQ and IPAQ surveys, we use intensity values from the WHO. The standard unit for PA intensity is the Metabolic Equivalent of Task (MET): the ratio of the energy spent on a task compared to the energy spent sitting quietly. In line with the recent DRFs meta-analyses (Garcia et al., 2023), we express PA intensity in marginal Metabolic Equivalent of Tasks (mMETs) rather than standard MET. We do this to capture the marginal energy expenditure that occurs due to adding PA to an individual’s daily routine. We assumed routine PA to be resting; thus, mMET is MET - 1 (Garcia et al., 2023). For example, the compendium PA values for transport-related cycling and walking are 6.8 and 3.5, leading to mMETs values of 5.8 and 2.5, respectively. An individual’s PA volume from any activity is the product of PA duration and intensity. So, an individual who cycled for one hour and walked for one hour during a survey week would have a transport-related PA volume of 8.3 (= 1x5.8 + 1x2.5) mMET-hours/week. The PA surveys capture different physical activities, their durations, and frequencies, which can be combined to calculate individual physical activity volumes. Further data processing and harmonisation involves extracting the non-occupational, non-transport PA component for one week. PA data are often outdated or lacking for many cities, prompting the use of surveys from contextually similar cities. In ITHIM-Global, we use two parameters to account for bias and uncertainty of background PA data, of which only one (`background_pa_scalar`) is relevant when running the model in the fixed input parameters mode (Table S14). For Bogotá, we judged the survey to adequately represent PA and set that parameter to 1, because the proportion of people achieving weekly WHO-recommended PA from moderate-to-vigorous activities in the survey was like estimates from Bogotá and other Latin American cities (Salvo et al. 2017).

The background non-transport, non-occupational PA component is assigned to individuals in the baseline population, as previously described in Section 2.2. Because the PA survey participants differ from those in the travel survey, the two datasets are combined probabilistically using age and sex to incorporate both transport-related and non-transport PA into the baseline population. We assume that non-transport, non-occupational PA is fixed across scenarios. Plausibly, an individual may exercise less if they commute more by cycling; alternatively, they may exercise more due to greater fitness (Foley et al., 2018). Inactive individuals with the lowest non-transport PA have the least PA to lose from any change in walking and cycling but the most to gain due to the non-linear DRFs (Garcia et al., 2023).

For Bogotá, we obtained background non-transport, non-occupational PA data from the PA module of the 2015 Colombian National Nutrition survey from the Ministry of Health (the most recent data available at the time of writing). The data contained weekly durations (minutes) of leisure walking and moderate and vigorous PA. Processing included transforming durations into hours and multiplying by 2.5, 3 and 7 mMETs, respectively.

Information for individual transport-related PA results from the walking and cycling trips in the travel survey. This PA component varies across scenarios as walking and cycling trips change. While PA surveys also contain transport-related PA, we do not use that domain because the travel surveys already contain detailed transport-related PA data, and the PA surveys are not at the trip level and hence do not allow the possibility of shifting trips, as does the travel survey. However, users should check the PA distribution in both surveys for verification. Total individual PA volumes for the baseline population in Bogotá are, therefore, the sum of non-transport, non-occupational PA (essentially leisure-time as domestic PA was not captured) from the 2015 PA survey and transport-related PA from the 2019 travel survey.

To model the health impacts of PA, the selection of DRFs is critical. Epidemiological studies have reported the effect of PA on many health outcomes. To estimate the impact of PA on all-cause and cause-specific mortalities and YLL, we used non-linear DRFs from recent meta-analyses (Garcia et al., 2023; Smith et al., 2016: for diabetes only). These DRFs are available as an R package and contain RR for 25 health outcomes^1^. ITHIM-Global uses only 17 of these health outcomes for which evidence was stronger, and health and PA gains are consistent. In most cases, there were diminishing marginal differences in RR and increasing uncertainty at high PA volumes, particularly >17.5 mMET-hours/week (Garcia et al., 2023); hence, we placed a cut-off at 17.5 mMET-hours/week for all outcomes, with no gains modelled from this censoring point onwards. PA health impacts are estimated at the three levels using these DRFs, as shown in Figure 3. Level 1 impacts refer to the highest or most inclusive level of outcomes, within which levels 2 and 3 fall. Level 1 is all-cause mortality, while level 2 includes high-level subtypes of mortality by organ systems, which for PA are cardiovascular diseases (CVD) and cancer.

Then, level 3 represents specific sub-outcomes within level 2 and includes 14 specific outcomes, as shown in Figure 3. Where a level 3 risk ratio did not have a corresponding level 2 cause, we included the level 3 risk ratio within level 2 by grouping these outcomes into the “Other” category. We selected risk ratios based on mortality when these were available. Still, for selected outcomes (Colon cancer, Uterine cancer, Liver cancer, Alzheimer’s disease and other dementias, Multiple myeloma, Parkinson’s disease, Head and neck cancer, Stomach cancer, Myeloid leukaemia), we only had sufficient evidence from studies that combined mortality and incidence, and diabetes mellitus type 2, we only had studies based on incidence, and assumed these were applicable to mortality.

Individuals’ RR are mapped from their PA volume at baseline and at a selected scenario and compared; thus, RR changes can be estimated at an individual level. However, since baseline rates for mortality and YLL are only available for fixed age bands by sex, we aggregate individual RR changes to match those bands. Potential impact fractions (PIF) are then calculated for each age group by sex and combined with baseline mortalities and YLL to obtain PA’s health impacts.

### 3.4. Module 3: Air Pollution Pathway

The air pollution module estimates particulate matter with less than 2.5 micrometres in diameter (PM_2.5_) and its attributable mortalities and YLL. PM_2.5_ was the pollutant included in ITHIM-Global as the datasets and information required to model PM_2.5_, its traffic contribution, and health impacts are available through open-access sources, and its adverse health effects are well-established through supporting epidemiology and toxicology. The air pollution module estimates two pollution metrics which are used in the health impact calculations for both active travellers and remaining population:

1. The new ambient PM_2.5_ concentrations for each modelled scenario in the form of a fixed value at the city level, which can be compared to baseline PM_2.5_, and
2. The individual PM_2.5_ dose for the baseline population, accounting for travel and non-travel activities, elevated on-road exposures while travelling, and different inhalation rates during activities of varying intensities.

The module can estimate other pollutants, such as nitrogen dioxide (NO_2_), if required input data are available. The elements and parameters of this module and how they connect are shown in Figure 6 and Table S15 and described next.

**Figure 6.**
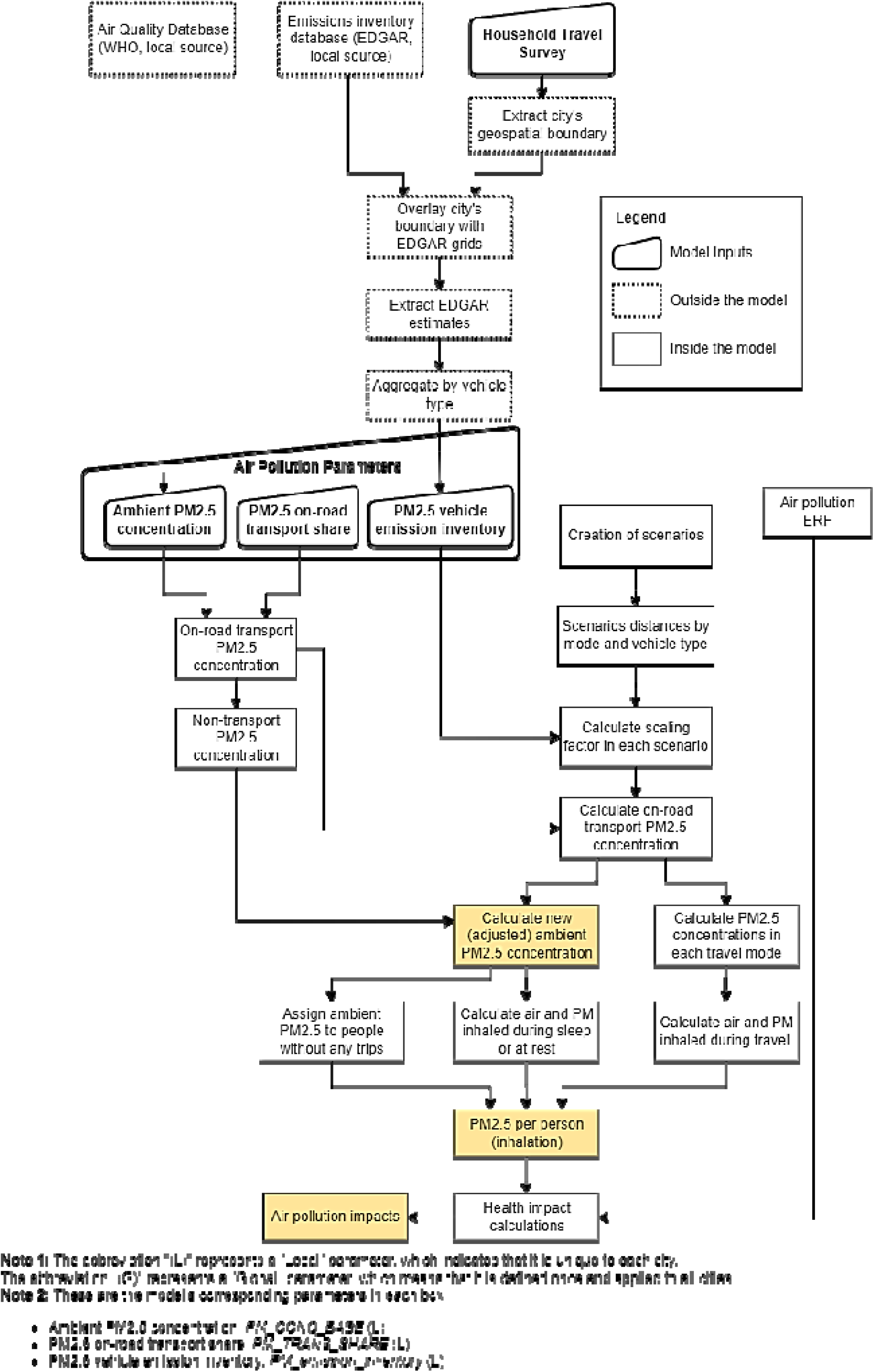
ITHIM-Global Workflow for the Air Pollution Module

The module first requires the ambient PM_2.5_ concentration (`pm_conc_base`), in the city of interest, which is treated as a fixed value applied across the city. No spatial variation of ambient PM_2.5_ is considered as we have no spatial information on where the baseline population lives and travels within the city. Ambient PM_2.5_ concentration can be obtained from regulatory air quality monitoring stations or systemised databases that often compile regulatory and other sources, such as WHO’s Air Quality Database. For Bogotá, we used the 2022 version of the database (Air Quality Database, 2022), which reported an average PM_2.5_ with 91% temporal coverage (Table S15). When presented with options, we recommend the use of urban background sites rather than rural, industrial, or roadside sites. An urban background is assumed to include road traffic contributions that are representative of the average exposure of the general population. If the population of interest is rural, industrial, or specifically lives near major roadways, then other air quality monitoring site types become relevant, and the source of ambient PM_2.5_ concentration for that population can be revised.

Next, we estimate how much of this value is from on-road transport sources using the percentage of PM_2.5_ due to on-road transport (`pm_trans_share`), which can be obtained from particulate matter source apportionment studies previously synthesised by WHO in Karagulian et al. (2015). Where data for the city of interest are unavailable, which was the case for Bogotá, we recommend using other published data or methods. Data from neighbouring cities may be considered for imputation as well, although this approach can carry additional inherent uncertainty due to differences that influence PM_2.5_ such as coastal versus inland terrain or meteorological factors. We used results from the Bayesian hierarchical model reported by Heydari et al. (2020). As our modelling is only concerned with passenger transport-related scenarios (i.e., changes to transport behaviours, mode shares, transport emissions, etc.), the PM_2.5_ concentration from non-transport sources was assumed to remain constant across scenarios. This is a limitation of our approach as changes in transport emissions may alter chemical regimes of ambient air and therefore formation of secondary particulate matter. A dispersion or chemical transport model can simulate these changes, but are data and expertise intensive, and time-consuming. However, users may alter non-transport sources PM_2.5_ based on simplified assumptions for more nuanced scenarios, e.g., to investigate how benefits of cycling increase if the ambient air is less polluted.

The third data input needed is the percentage (or total, which gets converted to percentage during an ITHIM-Global run) of PM_2.5_ vehicle emissions due to different vehicle types, i.e., the PM_2.5_ vehicle emissions inventory. A local vehicle emissions inventory using a bottom-up approach, which combines local vehicle-specific emission factors with local traffic activity and fleet distribution data, is preferred. However, this is more difficult to find or generate and a more readily available approach, which can be applied widely, was developed. We sourced PM_2.5_ vehicle emissions inventories from the Emissions Database for Global Atmospheric Research (EDGAR). EDGAR is a bottom-up model for estimating emissions, relying on a large spectrum of human activity data combined with emission factors to yield the emissions per source and country (Solazzo et al., 2021), with higher uncertainties in LMIC (Crippa et al., 2018). For the transport sector, EDGAR uses country-specific energy consumption statistics and fleet distributions obtained from the International Energy Agency (IEA), in addition to region-specific end-of-pipe/emission-exhaust-level abatement measures based on the more aggregate ’Tier 2’ methodology in EMEP/EEA, 2019. The calculated country-level emissions are scaled down to smaller geospatial grids, and EDGAR provides percentage PM_2.5_ emissions due to motorcycles/mopeds, cars, buses, and heavy and light duty trucks for 0.1-degree by 0.1-degree grids (≈ 11.1 by 11.1 km). EDGAR is likely less accurate and representative than national vehicle emission inventories, but it estimates transport emissions in a consistent and comparable way globally (Crippa et al., 2016).

To be used in ITHIM-Global, we spatially match the 0.1-degree by 0.1-degree PM_2.5_ emission grids to the geographical boundaries of the travel survey. This is done by overlaying the city’s geospatial boundary as defined in or inferred from the travel surveys with the EDGAR grids and extracting EDGAR estimates that fall within that boundary. We used the latest EDGARv6.1 at the time of analysis, from 2018, which included both exhaust and non-exhaust emissions.

The fourth step, done within an ITHIM run, is deriving a scaling factor (SF) as shown below:

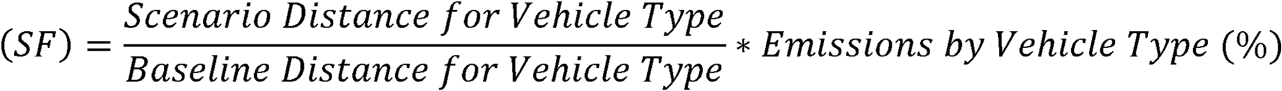

The emission percentage for the corresponding vehicle type were calculated from EDGAR. This process will estimate a new emission percentage for each vehicle type-scenario combination. As we are only interested in changes in passenger travel, heavy- and light-duty truck distances and their PM_2.5_ contribution remain unscaled. The scaling described offers a methodology applicable to LMIC in instances where air quality modelling is not possible or feasible. However, it is limited by the assumption that the relative distances used to compile EDGAR emissions (part of the activity data per vehicle type) are similar to the relative distances per vehicle type from our travel distance data, which may not be the case. This approach also assumes that we can align the definition of vehicle types between EDGAR and the travel survey. It then simplistically assumes that changes in distances by vehicle type equal changes in emissions (and subsequently PM_2.5_ concentrations), with no impact of chemistry or pollution from other sources. We multiply this scaling factor per vehicle type by the PM_2.5_ concentration in the city from on-road transport to estimate new PM_2.5_ concentration from on-road transport after scenario distances have changed. Finally, concentrations from non-transport sources are added to estimate a new ambient PM_2.5_ in each scenario (metric 1).

Two other datasets are needed to estimate metric 2: individual PM_2.5_ exposures. The first data are the mode-specific conversion factors to convert the new (scenario-adjusted) ambient PM_2.5_ concentration in the city to PM_2.5_ concentrations on the road experienced by different mode users. Indirectly, this accounts for the spatial variation in PM_2.5_ when travelling (i.e., when near or away from roadways). We used mode-specific conversion factors from WHO’s HEAT tool (Kahlmeier et al., 2017). These factors (Table S15) originate from a quantitative review aggregating data from European cities and are, therefore, not city-specific and may differ from LMICs through differences in, e.g., fleet mix and fuels (De Nazelle et al., 2017).

The second dataset is the inhalation rates per activity across all activities of the baseline population. To estimate the inhalation rate for each activity, we use a probabilistic approach that converts each activity and its corresponding MET value to an estimate of oxygen uptake, which in turn is transformed into an inhalation rate, following an algorithm in Johnson, (2002), and assumptions about time use by activity, discussed in the Section S13. The volume of air inhaled during each activity (m^3^) is then multiplied by the PM_2.5_ concentrations experienced to estimate the PM_2.5_ doses each individual is exposed to. These doses are then summed across all activities in the day and divided by air inhaled during the day to come up with individual exposures. The day’s exposure is assumed to represent the average annual exposure. The process above has been automated in the model^2^.

ERFs were selected from the published literature to estimate the impact of PM_2.5_ exposures on all-cause and cause-specific mortalities and YLL. As with PA, outcomes were categorised in a hierarchy where level 1 is all-cause mortality, under which levels 2 and 3 fall. For levels 1 and 2, we used ERFs to estimate the associations between long-term PM_2.5_ and all-cause and cause-specific mortalities from a random-effect meta-analysis conducted for the update of the WHO Global Air Quality Guidelines (Chen and Hoek, 2020). Level 2 outcomes included were: 1. circulatory diseases and 2. non-malignant respiratory diseases. This was the most recent at the time of writing, including 17-25 studies mostly from Europe,

Canada, and the United States and a final certainty assessment of high to moderate and an underlying PM range of 1 in Pinault et al. (2016) to 83.8 µg /m^3^ in Yin et al. (2017). This is a limitation in our approach as we rely on published ERFs, which currently include no or few studies in the African, Latin American, Southeast Asian, and Eastern Mediterranean Regions.

For level 3 outcomes, we used ERFs from the GBD 2019 Particulate Matter Risk Curves (Global Burden of Disease Collaborative Network, 2020). The risk curves have been estimated using cohort and case-control studies for ambient air pollution and second-hand smoking, in addition to the two study types, randomised control trials for household solid fuel use^3^. Among the selected six cause-specific mortality outcomes shown in Figure 3, the ERFs are age-specific for IHD and stroke, as RR for cardiovascular diseases has been found to decrease with age. For these two outcomes, RR were presented as lookup tables for each 5-year age band from 25-29, 30-34, and so on up to 95-99 (<25 RR was set to 1).

### 3.1. Module 4: Carbon Dioxide Emissions

ITHIM-Global models carbon dioxide (CO_2_) emissions from motorised vehicles. CO_2_ is the main greenhouse gas associated with road transport, representing about 99% of the climate change impact of road transport (IPCC, 2022). The datasets and information required to model traffic-related CO_2_ and its changes across modelled scenarios are available through open-access sources, as will be described in this section. The model estimates two metrics: total CO_2_ emissions and CO_2_ emissions per capita for each modelled scenario. As with air pollution, these scenario outputs are at the city level and compared to the baseline scenario. The method to estimate CO_2_ emissions is like the one for PM_2.5_, so we limit the description here to different aspects of how CO_2_ emissions are derived. Elements and parameters of this process, and how they feed back into each other, are shown in Figure 7.

**Figure 7.**
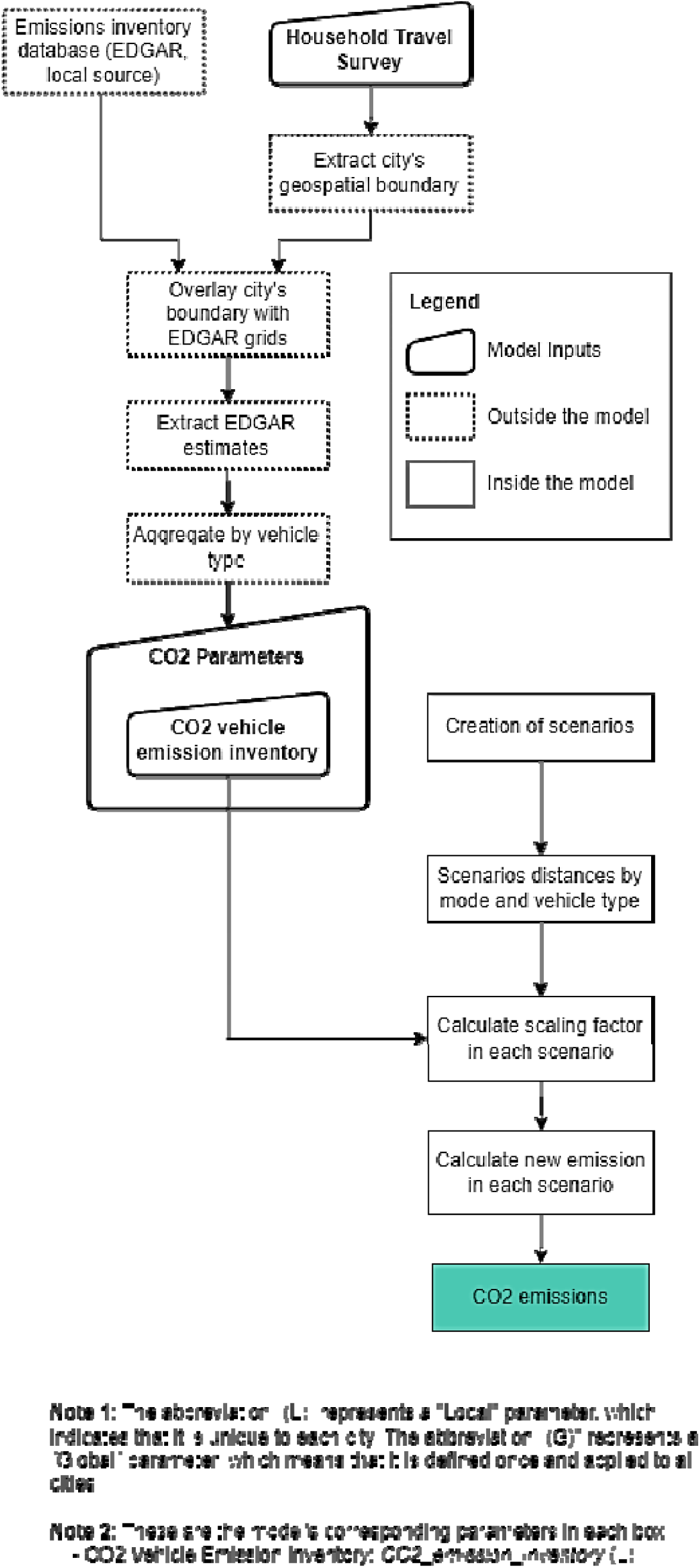
ITHIM-Global Workflow for CO2 Emissions Estimation

We sourced CO_2_ vehicle emissions from EDGAR v6.1 (Solazzo, Crippa et al. 2021). EDGAR ultimately provides total and percentage share CO_2_ emissions from motorcycles/mopeds, cars, buses, and heavy and light duty trucks at 0.1-degree x 0.1-degree grids (≈ 11.1 x 11.1 km), which we spatially matched to geographical boundaries of the travel surveys. After extracting all points within the boundary, total CO_2_ emissions can be summed by vehicle type, and then the percentage contribution of each type to the total emissions calculated (vehicle emissions inventory). This process has been automated in R^4^ and requires EDGAR CO_2_ emission geospatial maps per vehicle type and the city’s geospatial boundary.

We derived a scaling factor by first dividing the scenario distance by the baseline distance (for motorcycles/mopeds, cars, buses, and light and heavy-duty trucks) and then multiplying this value by the CO_2_ emission percentage for the corresponding vehicle type as calculated from EDGAR, as shown below:

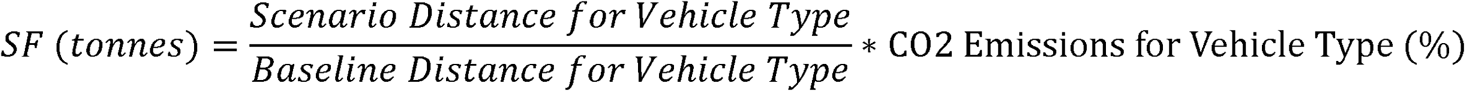

This produced a new emission percentage and total CO_2_ emissions (in tonnes) for each vehicle type- scenario combination. As we are only concerned with passenger travel (car, motorcycle/moped, cycling, bus), heavy and light-duty truck distances and contribution to CO_2_ remain unchanged across the scenarios.

The combination of EDGAR emission inventory estimates and the scaling methodology offers a consistent and comparable way applicable for LMICs. This approach is, however, limited by the same limitations outlined in the air pollution pathway Section (3.8). A further limitation concerns how the approach impacts small versus large cities differently. Smaller cities may have an interstate highway or motorway running through their territory, with associated CO_2_ emissions allocated to the city; yet residents of that city who may have completed the travel survey may not have travelled on those roadways, so it overestimates per capita emissions. For larger cities, this effect may average out.

## 4. RESULTS

### 4.1. Running the Model

We developed and piloted an open-access and transparent R package that runs the model described in this article. The code, required data, and documentation are on GitHub at: https://github.com/ITHIM/ITHIM-R/. In addition, we wrote a specific <u>guide</u> on how to run Bogotá’s ITHIM-Global in R, update inputs, and produce results of the modelled scenarios, where we detail:

- How to install the ITHIM-R package
- Running ITHIM-Global
- Results
- Structure of ITHIM-Global GitHub repository - Bogota branch

### 4.2. Generating and Navigating Outputs

Running the model results in many outputs. We broadly classify these into two types:

1. <u>Intermediate outputs,</u> which users can examine to check the model’s sensibility, verify intermediate steps, deepen their understanding and interpretation of processes and results, and pinpoint any discrepancies and errors in the modelling pipeline,
2. <u>Final outputs,</u> which are the modelled mortalities and YLL in each scenario.

In Section S14 (“Checking Intermediate Model Outputs”), we orient users through selected intermediate outputs and provide instruction on how to check their values and logic, interpret them, verify intermediate processes, and utilise them to understand and explain final outputs. The final outputs are in Figure 8 to Figure 16. They are also hosted interactively on Shiny at https://ithim-apps.shinyapps.io/results_app/.

**Figure 8.**
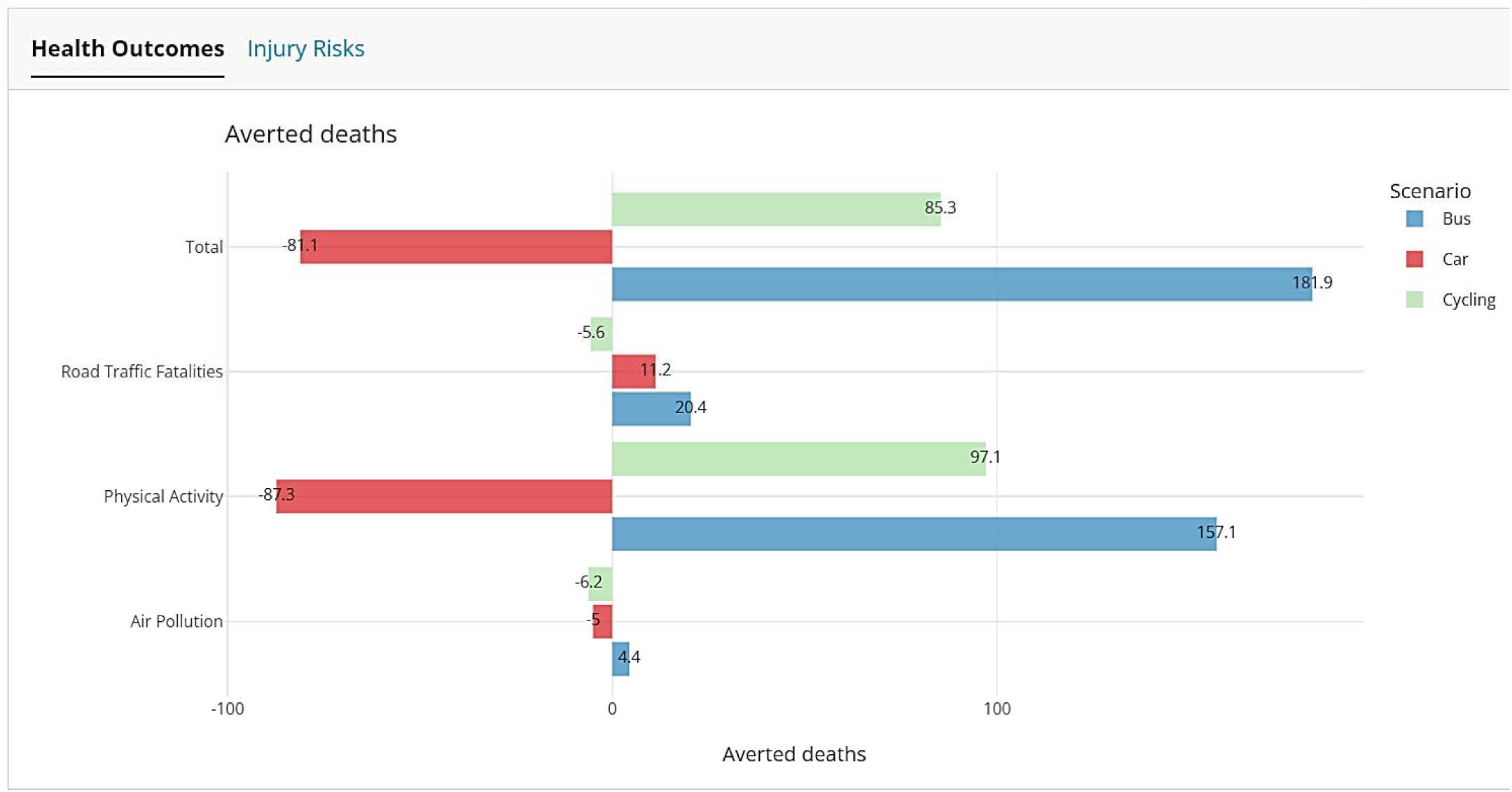
Mortality Averted or Attributable to the Three Scenarios for Level 1 Outcomes (All-Cause Mortality and Road Traffic Fatalities; See Levels in Figure 3

**Figure 9.**
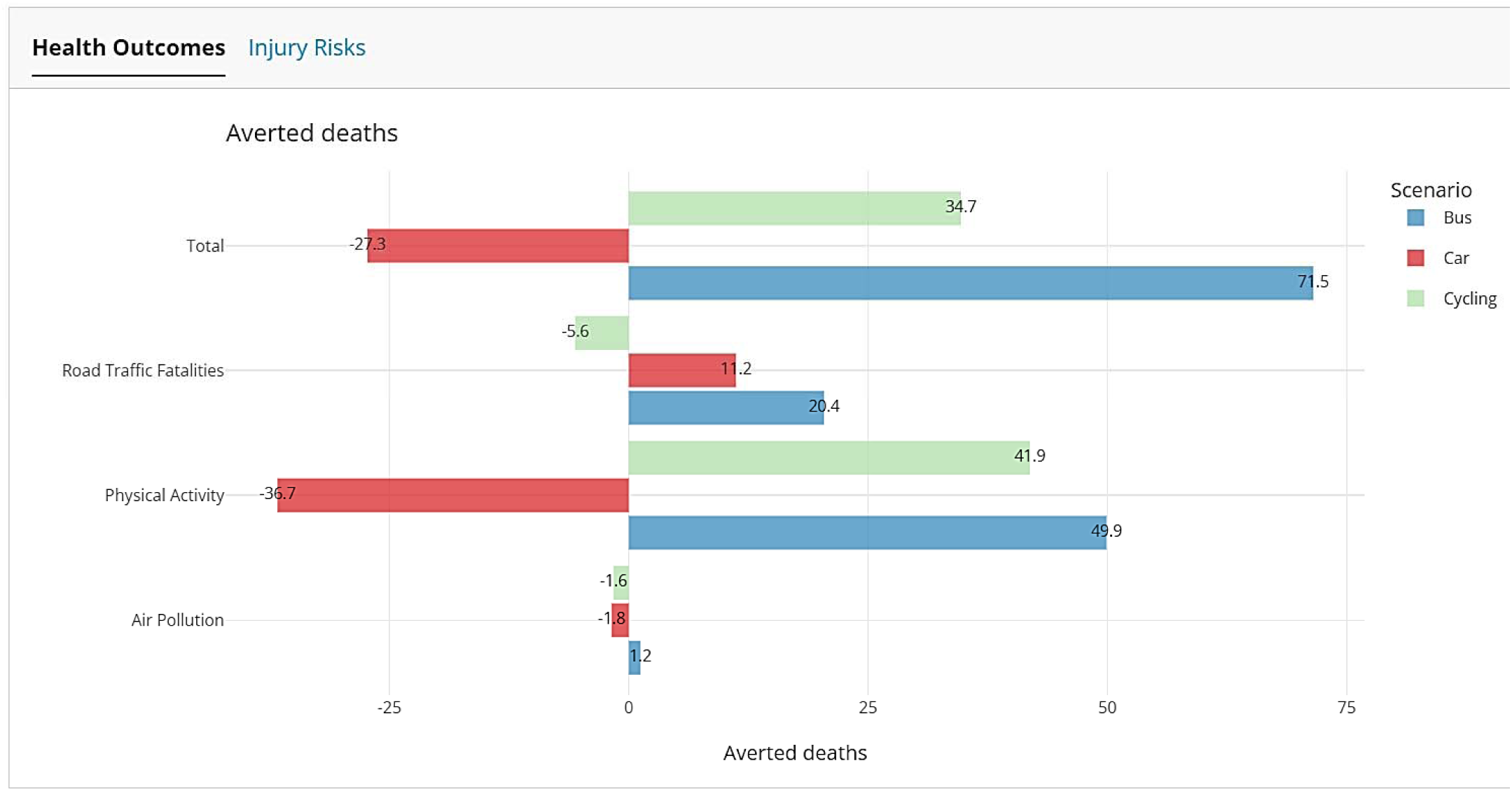
Mortality Averted or Attributable to the Three Scenarios for Level 2 Outcomes (Respiratory Diseases, Cardiovascular Diseases, Cancers, Others, Road Traffic Fatalities; See Levels in Figure 3

#### 4.2.1. Mortality by Pathway and Scenario Combinations for the Three Outcome Levels

The first set of results show averted (positive values) or attributable (negative values) mortalities by each pathway and scenario combination, in addition to total mortality from all combined pathways. The user can investigate mortality by outcome level: 1, 2 or 3. As shown in Figure 8 through Figure 10, the choice of the outcome level impacts the magnitude of averted/attributable mortalities. However, trends across pathway and scenario combinations are consistent: The bus scenario is the most beneficial as it averts mortalities across all pathways, primarily driven by increases in physical activity, while air pollution exerts the least influence. In the bus scenario, benefits from the PA pathway result from the shift from car trips and the inclusion of short walks to and from bus stops (see Sections S14.2 and S14.3). The cycling scenario is the second most beneficial, also primarily driven by increases in physical activity, while its harms are driven by increases in road traffic fatalities and mortality attributable to air pollution. As shown in Section S14.2, most of the trips in the cycling scenario come from previous bus and pedestrian trips. When cycling, a cyclist will have an increased inhalation rate while experiencing higher roadside PM_2.5_ concentrations and ultimately individual PM_2.5_ exposures increase for the active travellers, although it reduces the overall ambient PM_2.5_ concentration (see Section S14.4). While cycling is inherently a form of active travel, PA benefits are greater in the bus scenarios due to greater gains in PA among people with low baseline activity levels. In contrast, in the cycling scenario, the total increase in PA volume is greater, but more of it occurs at higher PA levels, specifically over the censoring threshold of 17.5 mMET-hours/week (see Section S14.3). In addition, some walking to and from bus stops and pedestrian trips are lost in the cycling scenario (see Section S14.2). Combined with the non-linear DRF and the censoring of benefits at 17.5 mMET-hours/week, marginal benefits are smaller at higher PA in the cycling scenario when compared to the bus scenario. The car scenario is adverse overall, with most trips coming from previous bus trips (See Section 14.2), resulting in large reductions in PA (see Section S14.3). the car scenario also increases air pollution exposure slightly, for both active travellers and the rest of the population (see Section S14.4), while it yields small benefits from a reduction in road traffic fatalities (see Section 14.5).

**Figure 10.**
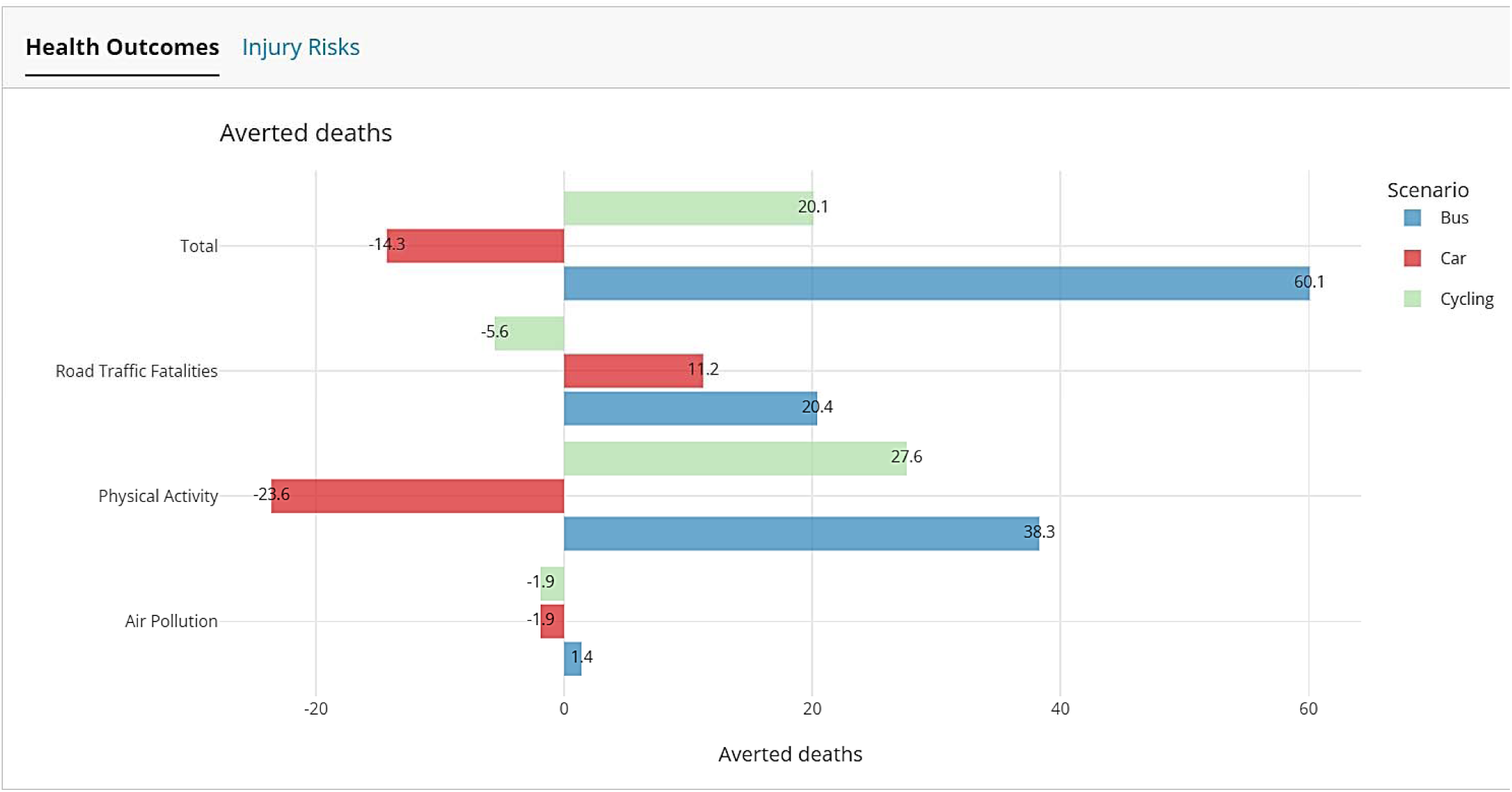
Mortality Averted or Attributable to the Three Scenarios for Level 3 Outcomes (Specific Diseases e.g., COPD, Stroke, Type 2 Diabetes and Road Traffic Fatalities; See Levels in Figure 3

For all pathways, the magnitude of impacts is largest using level 1 outcomes (total averted mortalities = 182 in the bus and 85 in the cycling scenario, while total attributable mortalities = 81 in the car scenario), and smallest for level 3 outcomes (total averted mortalities = 60 in the bus and 20 in the cycling scenario, total attributable mortalities = 14 in the car scenario).

The user can also investigate the impact of an interaction between air pollution and physical activit (Figure 11), and stratify by sex (Figure 12) and age (Figure 13) to explore the distribution of impacts amongst subgroups. For example, Figure 12 shows more deaths averted among males than females through the physical activity pathway in the bus and cycling scenarios. This is sensible because both inactive and active males at baseline gained more physical activity than females in those scenarios, whereas similar proportions of males and females had low baseline physical activity. Users can check this by exploring the distribution of physical activity output.

**Figure 11.**
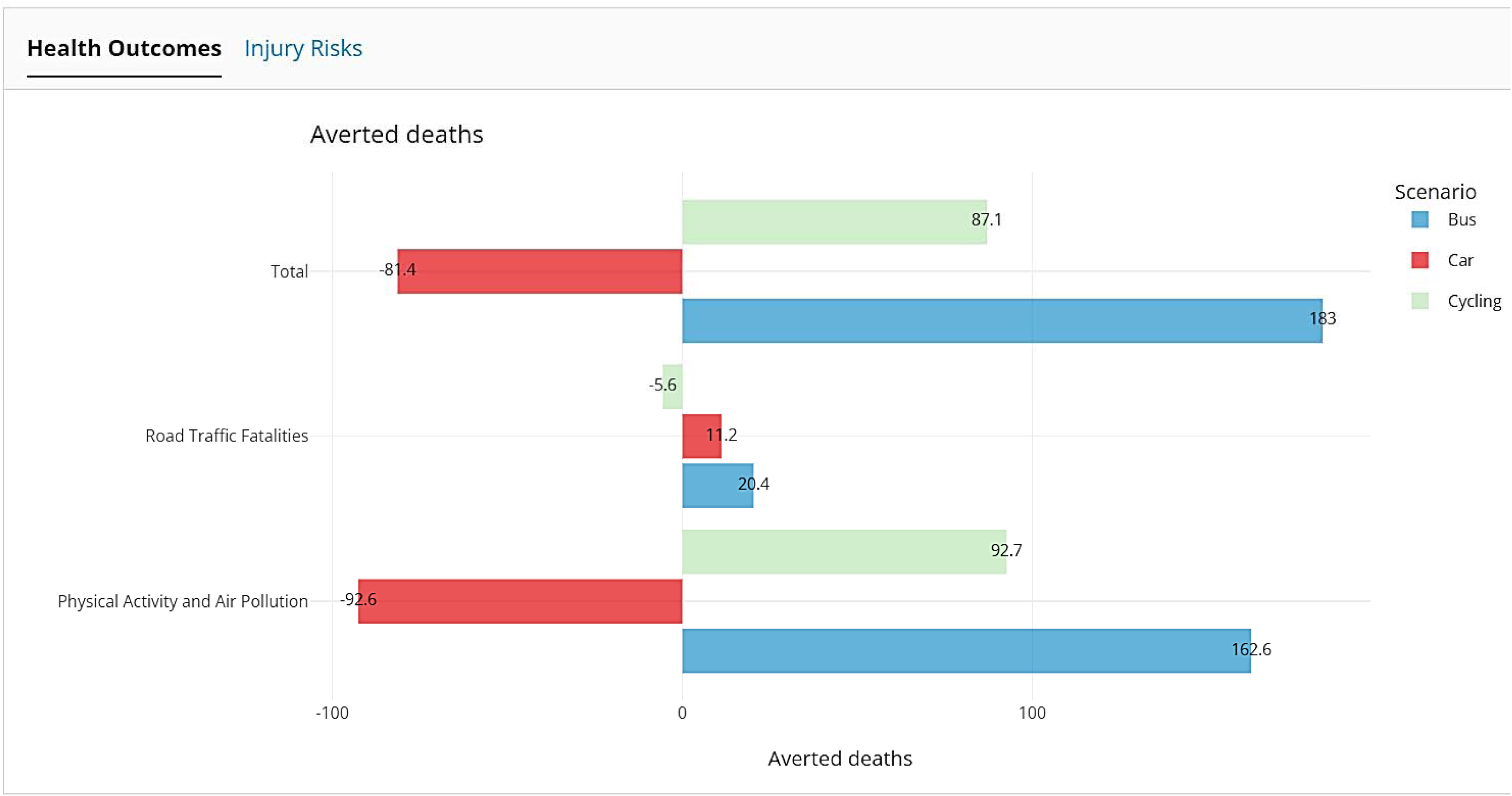
Mortality Averted or Attributable to the Three Scenarios for Level 1 Outcomes (All-Cause Mortality and Road Traffic Fatalities; See Levels in Figure 3) including an Interaction Between Physical Activity and Air Pollution

**Figure 12.**
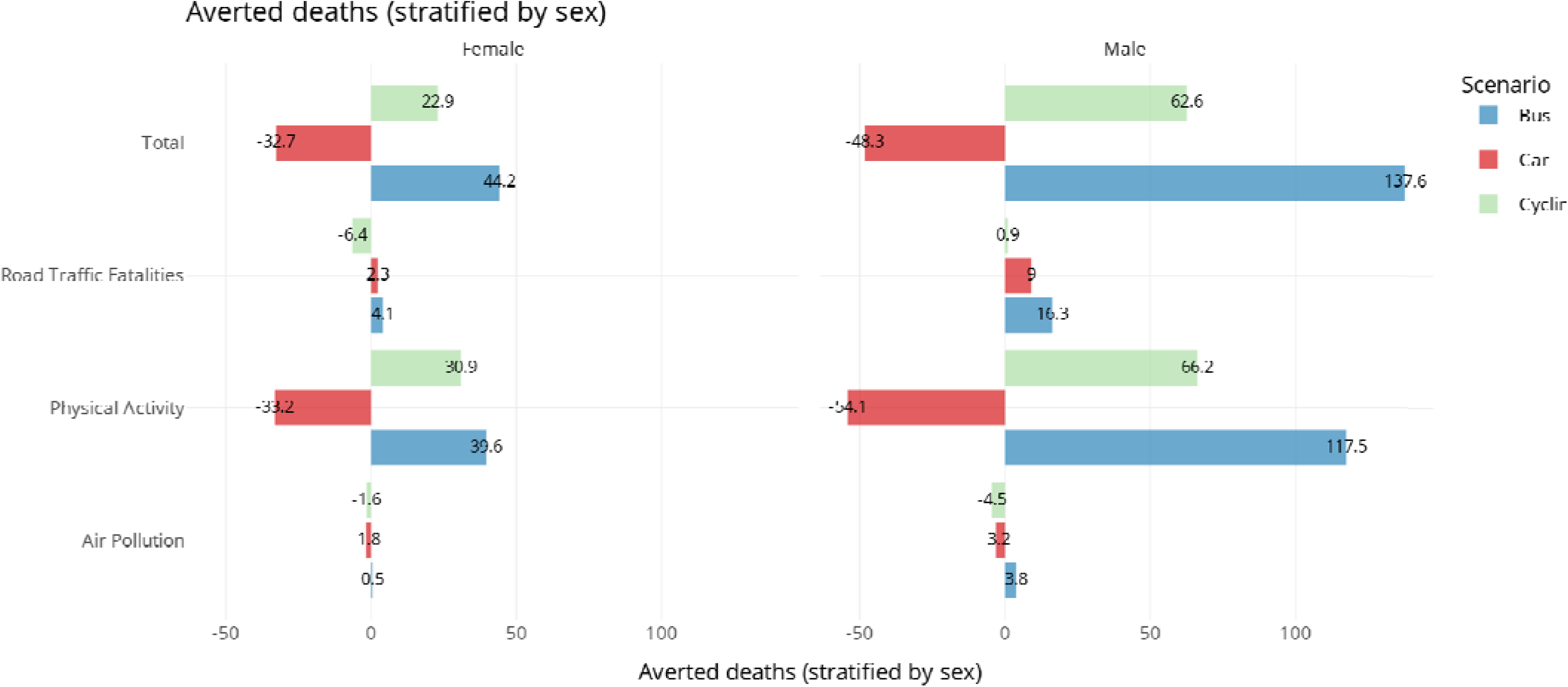
Mortality Averted or Attributable to the Three Scenarios for Level 1 Outcomes (All-Cause Mortality and Road Traffic Fatalities; See Levels In Figure 3) Stratified by Sex

**Figure 13.**
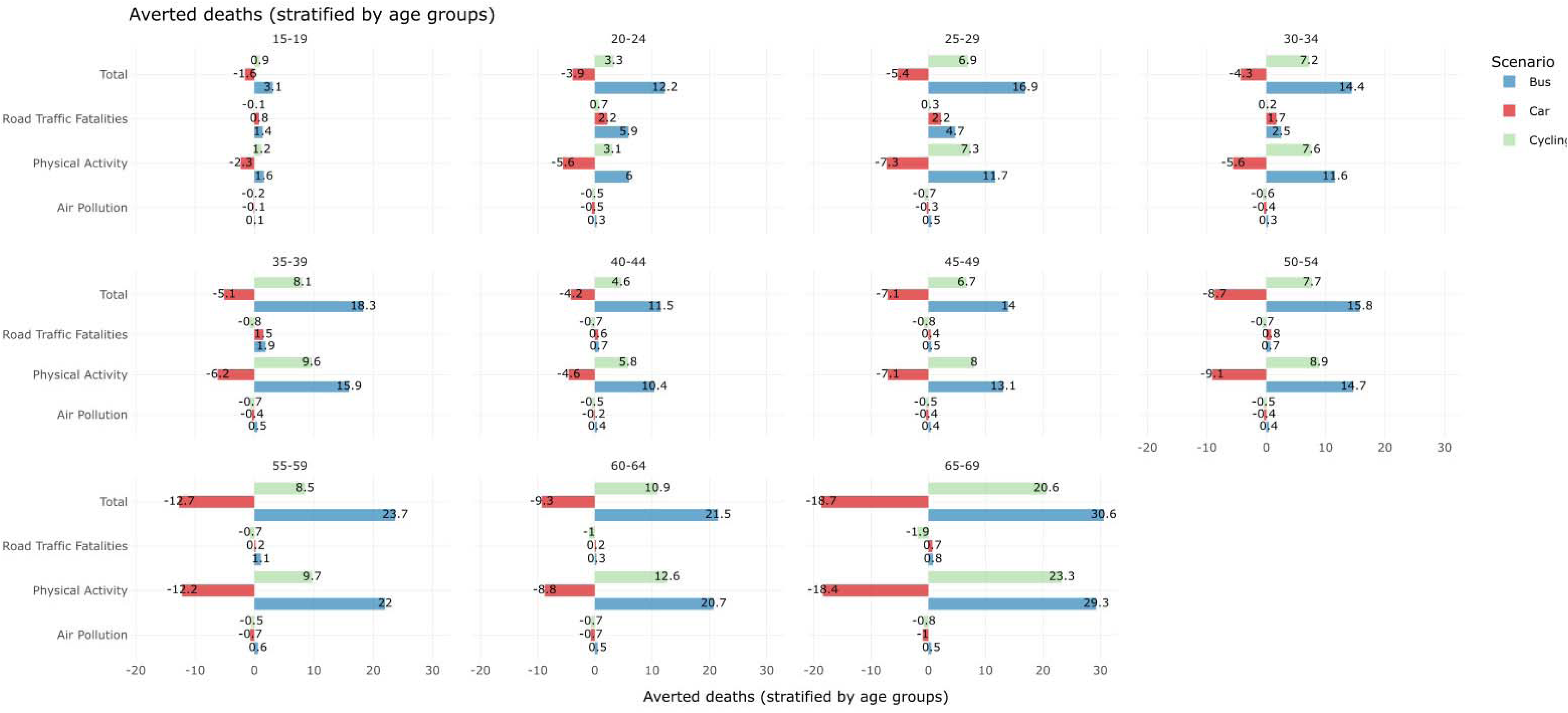
Mortality Averted or Attributable to the Three Scenarios for Level 1 Outcomes (All-Cause Mortality and Road Traffic Fatalities; See Levels In Figure 3) Stratified by Age 50

#### 4.2.2. YLL by Pathway and Scenario Combinations for the three Outcome Levels

The second set of results are the averted or attributable YLL by each pathway and scenario combination, in addition to the total YLL from all pathways combined. The user can investigate these similarly to the mortality outputs (e.g., Figure 14).

**Figure 14.**
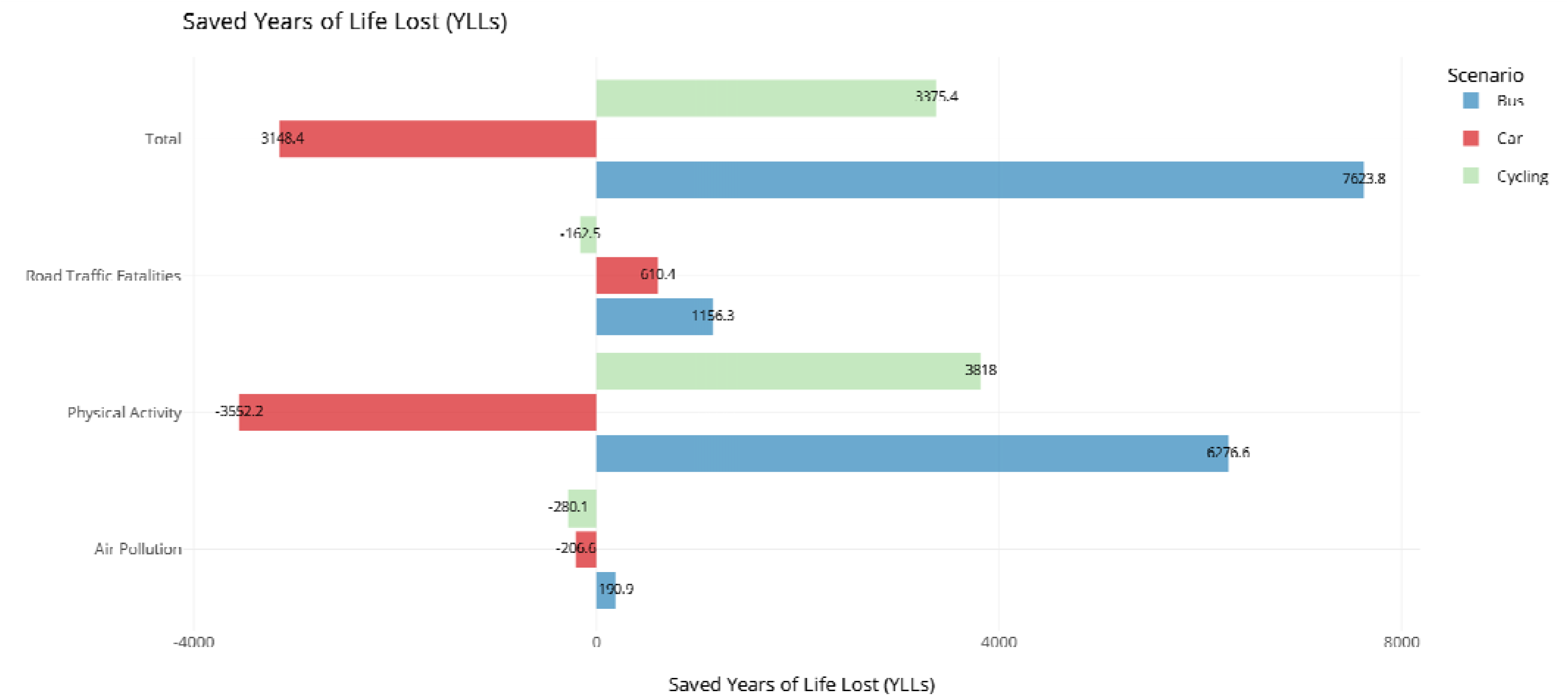
Years of Life Lost Averted or Attributable to the Three Scenarios for Level 1 Outcomes as an Example (All-Cause Mortality and Road Traffic Fatalities; See Levels in Figure 3)

#### 4.2.3. Carbon Dioxide Emissions

Table 2 shows the CO_2_ emissions in the baseline and how it changes across the three scenarios. The bus scenario emerges as the most beneficial followed by the cycling scenario, while the car scenario increases CO_2_ emissions by 2.2%. This is due to different trip reassignments for each of the scenarios, as explained in Section S14.2.

**Table 2.** Carbon Dioxide Emissions in the Baseline and the Three Created Scenarios in Tonnes.

| Scenario | CO<sub>2</sub> Emissions (tonnes) | Change from Baseline | Percentage Change from Baseline |
| --- | --- | --- | --- |
| Baseline | 1,852,649 | NA | NA |
| Bus | 1,827,352 | -25,297.3 | -1.4 |
| Car | 1,894,025 | 41,375.8 | 2.2 |
| Cycling | 1,836,973 | -15,676.3 | -0.8 |

#### 4.2.4. Sensitivity Analysis Results

The results from sensitivity analyses are shown in Figure 15 and Figure 16 for the 1% and 10% scenarios. Trends across pathway and scenario combinations are consistent with the main analysis (5%) with proportional increases in averted or attributable deaths.

**Figure 15.**
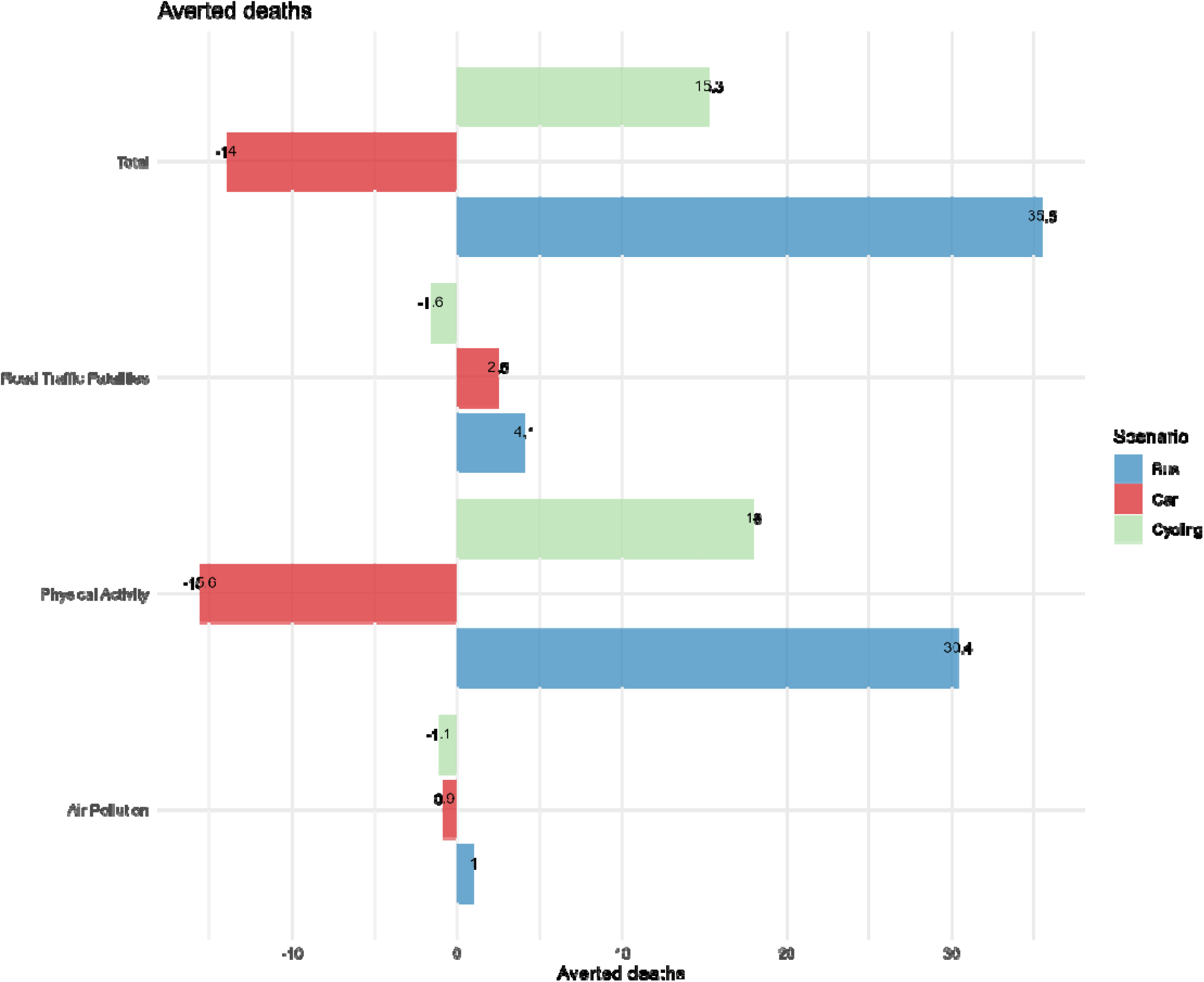
Mortality Averted or Attributable to the Three Scenarios for Level 1 Outcomes (All-Cause Mortality and Road Traffic Fatalities; See Levels in Figure 3Error! Reference source not found.) using a 1% Increase in Target Modes

**Figure 16.**
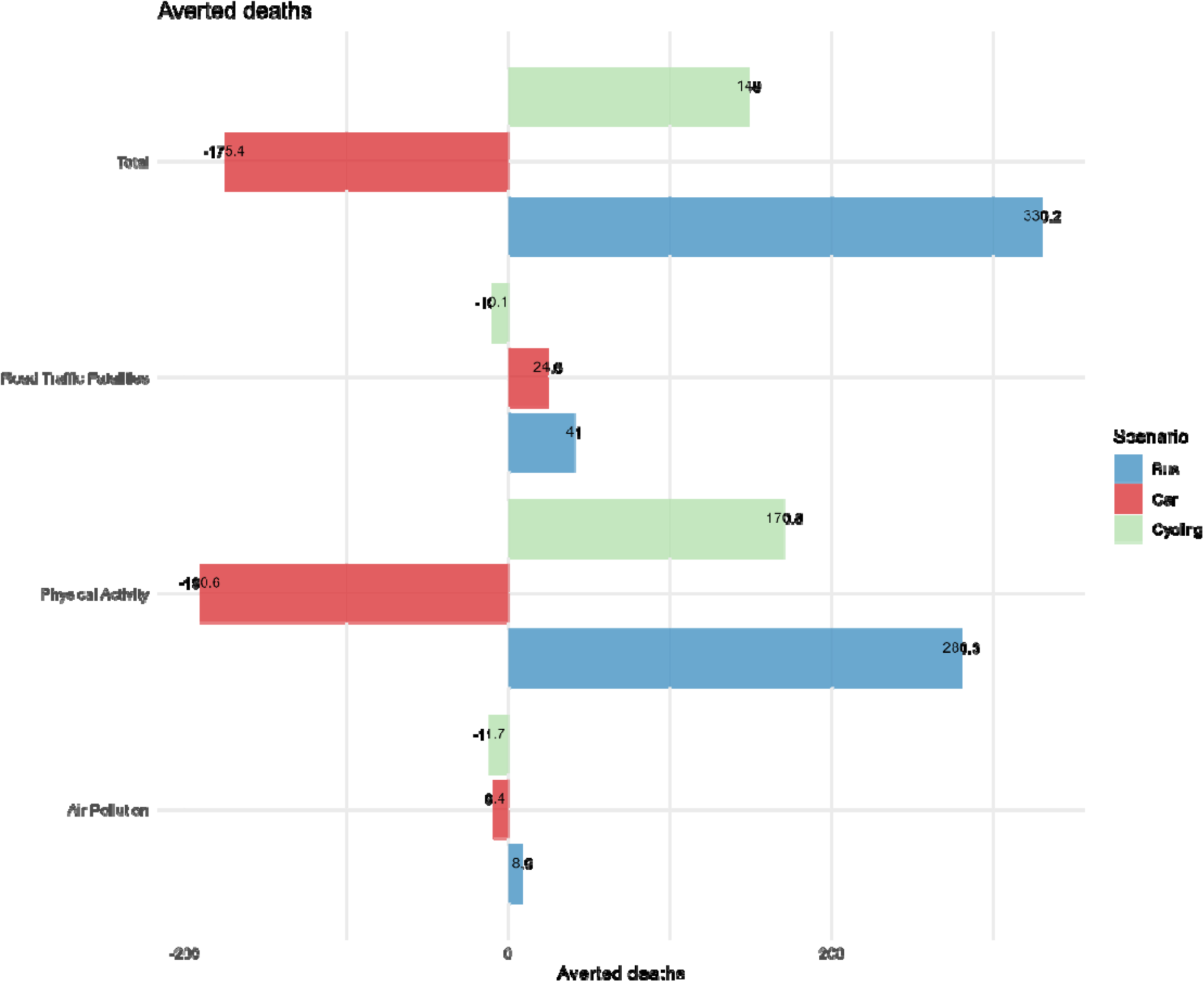
Mortality Averted or Attributable to the Three Scenarios for Level 1 Outcomes (All-Cause Mortality and Road Traffic Fatalities; See Levels in Figure 3) using a 10% Increase in Target Modes

## 5. DISCUSSION

### 5.1. Summary

We developed the ITHIM-Global model for HIA of transport in LMICs and showcased its application by simulating a 5% mode shift in Bogotá across three target modes. ITHIM is a widely used HIA model in transport studies, followed by the WHO’s HEAT tool, and we compare the two in Section 5.3 (Mizdrak et al., 2023). We outlined the model’s assumptions and their influence on outputs by examining and verifying intermediate outputs, in much detail in the Supplementary Material. This thorough evaluation of inputs and outputs empowers users to leverage the outputs for policymaking, while addressing contingencies and context-specific factors captured by the model.

### 5.2. Strengths and Limitations

ITHIM-Global was specifically designed to use data readily available in LMICs. We detailed the data sources, quality control processes, assessments, and ideal alternatives. Together with this article, the model can support the adoption, further development, and expansion of ITHIM-Global and similar models in LMIC cities, where such HIA are urgently needed. Key advancements generalisable for HIA include capturing individual data from travel surveys, assigning personalized exposure estimates, and assessing multiple health pathways and their interactions—such as disaggregating inhalation rates by age, sex, and physical activity and including an interaction term between air pollution and physical activity.

ITHIM-Global is not designed to simulate health outcomes in children and currently focuses on mortality and YLL. While physical activity in children is linked to developmental and mental health benefits (Biddle et al., 2019), it is not strongly associated with mortality (Chaput et al., 2020). Although early-life physical activity may influence activity levels later in life, incorporating this into a static comparative risk assessment model introduces considerable uncertainty, including in discounting future benefits. Additionally, the model does not account for occasional childhood mortality due to air pollution, which primarily affects development and disease outcomes. Traffic fatalities occur at all ages. However, focusing solely on road traffic fatalities and air pollution-related mortality in children, while excluding the health potentially large benefits of physical activity, would introduce bias.

Future assessments should include morbidity impacts, particularly in children, and consider non-fatal traffic injuries, which are often underreported far more than fatal crashes. Including older adults in assessments typically results in larger health gains from reductions in air pollution and increases in physical activity due to higher absolute mortality risk in this group and physical activity tending to be lower at older age. Although the YLL impact is smaller than mortality, it remains substantial. Using risk ratios based on studies of middle-aged individuals may also overestimate the benefits of increased physical activity in older adults. Finally, future models should explore and integrate occupational and objective physical activity data, the former often poorly captured in surveys. This is especially relevant in LMICs where occupational physical activity may be higher.

ITHIM-Global’s air pollution assessment is currently non-spatial but indirectly accounts for exposure variations in transport environments. It can be improved by integrating spatially resolved pollution maps with baseline population residence data (available in some travel surveys) and potentially work locations, alongside transport routing (e.g., shortest distance between origin and destination). We recommend including traffic-specific pollutants, such as black carbon and NO_2_ in future iterations, which are more sensitive to transport changes and mode shift scenarios, although data on their source apportionment and concentrations are less available than PM_2.5_. Future iterations of ITHIM-Global may address these data limitations using indirect methods such as those we developed in Navaratnam et al. (2024).

Another limitation is the reliance on ERFs that lack representation from Africa, Latin America, South-East Asia, and the Eastern Mediterranean, where dust may influence PM_2.5_ levels, making current risk estimates less applicable. Additionally, the model’s inhalation rates are based on outdated U.S. data with small sample sizes and need updating with data from LMICs. Time-use surveys, currently based on European data, should be adapted to reflect local surveys and account for subgroup variations previously recorded across regions, gender, and socio-economic factors (Rubiano et al., 2019; Medina- Hernández et al., 2021). Finally, the model’s use of one-day travel surveys limits its ability to differentiate between weekday and weekend travel patterns and does not account for seasonal changes in traffic or activities.

### 5.3. Comparison to HEAT

The HEAT model also evolved since launch in 2008, with version 5.0 designed for global applications. Initially focused only on physical activity, HEAT now includes air pollution exposure and injury risk, but only for active travellers, unlike ITHIM-Global, which assesses the entire population including active travellers. ITHIM-Global has therefore additional air pollution and road traffic fatality risk modules that HEAT does not require. HEAT 5.0 uses a log-linear DRF with a threshold between walking, cycling (considered separately), and all-cause mortality, while ITHIM-Global employs non-linear DRFs for all-cause and a wide range of cause-specific mortalities. As a result, HEAT can use mean exposure as a reasonable estimate, whereas ITHIM-Global uses a quasi-microsimulation approach to account for population variability and maximize the use of non-linear DRFs.

HEAT restricts the age range of the population and assumes average mortality rates for the age ranges of 20–74 years for walking and 20–64 years for cycling (or 20–74 years for e-biking) and should not be applied to populations of children or adolescents, since the underlying scientific evidence does not include these age groups (Health Economic Assessment Tool, 2024). In contrast, ITHIM-Global specifies ages and uses underlying mortality and YLL burden data in 5-year increments. HEAT offers a free web- tool, though it is not open source and is subject to WHO approval, while ITHIM-Global is open source, but currently lacks a web-tool, though an interactive visualizations on Shiny at https://ithim-apps.shinyapps.io/results_app/ have been created. HEAT primarily targets transport practitioners but is also widely used in academic research, offering assessments at both the city-wide and project levels. In contrast, ITHIM-Global focuses on city-wide assessments.

HEAT assumes constant inhalation rates across cities without accounting for variations in activity intensity, such as due to age and sex or cycling at different speeds or on gradients. ITHIM-Global, however, disaggregates inhalation rates by age, sex, and PA levels although it also does not account for speed and gradient variations. In brief, HEAT is intended to simulate an increase in the number of pedestrians or cyclists and the health outcomes for that population, while ITHIM-Global is intended to simulate health outcomes for changes to a city’s transport system which could be used without any changes in walking or cycling. Both HEAT and ITHIM have benefited from developments due to some commonality of people, with e.g. versions of ITHIM presented at HEAT meetings.

## 6. CONCLUSIONS

ITHIM-Global has been developed to facilitate HIAs of transport mode shifts in cities of LMICs. This model integrates multiple pathways, including physical activity, air pollution exposure, road traffic fatalities, and CO_2_ emissions. Applying the model to Bogotá, Colombia, revealed significant potential health benefits from a 5% shift in transport modes toward bus and cycling, primarily due to increased physical activity and reduced emissions. Such shifts also presented minor risks, such as increased exposure to air pollution and the risk of road fatalities for active travellers. The results underscore the importance of understanding context-specific trade-offs and benefits in transport policymaking. We identified areas for improvement in future iterations of ITHIM-Global, such as introducing spatially refined air pollution models, including children and the elderly, updating DRF and ERF, and capturing temporal variations and a wider array of health outcomes. These refinements will bolster the tool’s utility for policymakers aiming to craft transport strategies that optimize health outcomes.

## Data Availability

We obtained essential datasets to demonstrate the implementation of ITHIM-Global in Bogota from public domains and government agencies. Population data comes from Population projections disaggregated by locality 2018-2035 based on the National Population and Housing Census 2018 (https://www.dane.gov.co/index.php/estadisticas-por-tema/demografia-y-poblacion/proyecciones-de-poblacion/proyecciones-de-poblacion-bogota), travel behaviour comes from the 2019 Mobility survey from the Integrated Information System on Regional Urban Mobility (https://www.simur.gov.co/encuestas-de-movilidad), physical activity from the 2015 Colombian National Nutrition survey from the Ministry of Health (accessible upon request), road traffic injuries from the police department shared by the Bogota Secretary of Mobility (https://datos.movilidadbogota.gov.co/search?groupIds=d3812f8315054cdc84cf744680103713), air pollution data from the 2022 version of WHO Air Quality Database (https://www.who.int/data/gho/data/themes/air-pollution/who-air-quality-database), vehicle emissions from EDGAR shared by Monica Crippa from the European Commission Joint Research Centre, and baseline mortality and years of life lost data from the Global Burden of Disease study (GBD) published by the Institute for Health Metrics and Evaluation (https://ghdx.healthdata.org/gbd-2019).

## Author Contributions

Conceptualization: HK, JW, AA; Data curation: HK, AA, DGS, LT, AS; Formal analysis: HK, AA, DGS, LT, AS, CB; Funding acquisition: JW; Investigation: HK, AA, DGS, LT, AS, RG, CB, JW; Methodology: HK, AA, DGS, LT, AS, RG, RJ, JW; Project administration: HK, AA, JW; Resources: HK, AA, DGS, LT, AS; Software: AA, AS, RJ; Supervision: HK, JW; Validation: HK, AA, DGS, LT, AS, RG, CB, JW; Visualization: HK, AA, DGS; Roles/Writing - original draft: HK; and Writing - review & editing: HK, AA, DGS, LT, AS, RG, CB, RJ, JW.

## Funding

Contributions by RG and JW were partially supported by TIGTHAT, an MRC Global Challenges Project MR/P024408/1. This project (JW, RG, LT, AA, AS, HK, DGS, CB) has received funding from the European Research Council (ERC) under the Horizon 2020 research and innovation programme (grant agreement No 817754). This material reflects only the author’s views, and the Commission is not liable for any use that may be made of the information contained therein.

### Acknowledgement

We gratefully acknowledge the contribution of the following colleagues: Kavi Bhalla from the University of Chicago’s Harris School of Public Policy for his advice on and provision of some injury data, Marko Tainio from the Finnish Environment Institute and Leandro Garcia from the Centre for Public Health at Queen’s University Belfast for previous related methods development, Christopher Jackson from the University of Cambridge’s MRC Biostatistics Unit for supervision of the statistical methods development and Monica Crippa from the European Commission’s Joint Research Centre for provision of vehicle emission inventories developed for the Emissions Database for Global Atmospheric Research (EDGAR).

## Data

We obtained essential datasets to demonstrate the implementation of ITHIM-Global in Bogotá from public domains and government agencies. Population data comes from “Population projections disaggregated by locality 2018-2035” based on the National Population and Housing Census 2018 (https://www.dane.gov.co/index.php/estadisticas-por-tema/demografia-y-poblacion/proyecciones-de-poblacion/proyecciones-de-poblacion-bogota), travel behaviour comes from the 2019 Mobility survey from the Integrated Information System on Regional Urban Mobility (https://www.simur.gov.co/encuestas-de-movilidad), physical activity from the 2015 Colombian National Nutrition survey from the Ministry of Health (accessible upon request), road traffic injuries from the police department shared by the Bogotá Secretary of Mobility (https://datos.movilidadbogota.gov.co/search?groupIds=d3812f8315054cdc84cf744680103713), air pollution data from the 2022 version of WHO Air Quality Database (https://www.who.int/data/gho/data/themes/air-pollution/who-air-quality-database), vehicle emissions from EDGAR shared by Monica Crippa from the European Commission’s Joint Research Centre, and baseline mortality and years of life lost data from the Global Burden of Disease study (GBD) published by the Institute for Health Metrics and Evaluation (https://ghdx.healthdata.org/gbd-2019).

## Footnotes

1 https://shiny.mrc-epid.cam.ac.uk/meta-analyses-physical-activity/

2 https://github.com/ITHIM/ITHIM-R/blob/bogota/R/scenario_pm_calculations.R

3 http://ghdx.healthdata.org/record/ihme-data/global-burden-disease-study-2019-gbd-2019-particulate-matter-risk-curves

4 https://github.com/ITHIM/CO2_EDGAR_2018/blob/main/script/co2_transport_emissions_edgar_2018.R

